# Clinical, Environmental, and Genetic Risk Factors for Substance Use Disorders: Characterizing Combined Effects across Multiple Cohorts

**DOI:** 10.1101/2022.01.27.22269750

**Authors:** Peter B. Barr, Morgan N. Driver, Sally I-Chun Kuo, Mallory Stephenson, Fazil Aliev, Richard Karlsson Linnér, Jesse Marks, Andrey P. Anokhin, Kathleen Bucholz, Grace Chan, Howard J. Edenberg, Alexis C. Edwards, Meredith W. Francis, Dana B. Hancock, K. Paige Harden, Chella Kamarajan, Jaakko Kaprio, Sivan Kinreich, John Kramer, Samuel Kuperman, Antti Latvala, Jacquelyn L. Meyers, Abraham A. Palmer, Martin H. Plawecki, Bernice Porjesz, Richard J. Rose, Marc A. Schuckit, Jessica E. Salvatore, Danielle M. Dick

## Abstract

Substance use disorders (SUDs) incur serious social and personal costs. Risk for SUDs is complex, ranging from social conditions to individual genetic variation. We examined whether models that include a clinical/environmental risk index (CERI) and polygenic scores (PGS) are able to identify individuals at increased risk of SUD in young adulthood across four longitudinal cohorts for a combined sample of N = 15,134. Our analyses included participants of European (N_EUR_ = 12,659) and African (N_AFR_ = 2,475) ancestries. SUD outcomes included: 1) alcohol dependence, 2) nicotine dependence; 3) drug dependence, and 4) any substance dependence. In the models containing the PGS and CERI, the CERI was associated with all three outcomes (ORs = 1.37 – 1.67). PGS for problematic alcohol use, externalizing, and smoking quantity were associated with alcohol dependence, drug dependence, and nicotine dependence, respectively (OR = 1.11 – 1.33). PGS for problematic alcohol use and externalizing were also associated with any substance dependence (ORs = 1.09 – 1.18). The full model explained 6% - 13% of the variance in SUDs. Those in the top 10% of CERI and PGS had relative risk ratios of 3.86 - 8.04 for each SUD relative to the bottom 90%. Overall, the combined measures of clinical, environmental, and genetic risk demonstrated modest ability to distinguish between affected and unaffected individuals in young adulthood. PGS were significant but added little in addition to the clinical/environmental risk index. Results from our analysis demonstrate there is still considerable work to be done before tools such as these are ready for clinical applications.

## INTRODUCTION

Substance use disorders (SUDs) are associated with substantial costs to affected individuals, their families, and society. An estimated 107,000 Americans died as the result of an overdose in 2021 ^1^. In 2016, alcohol use contributed 4.2% to the global disease burden and other drug use contributed 1.3% ^2^. Excessive alcohol use and illicit drug use cost the United States an annual $250 billion ^3^ and $190 billion ^4^ respectively. Given the substantial human and economic costs of substance misuse and disorders, understanding the combined impact of important risk factors across multiple levels of analysis has important public health implications.

Substance use disorders are complex phenomena, and the development of substance related problems can be attributed to factors ranging from broader social and economic conditions to individual genetic variation ^5–10^. Prior research using a multifactorial index of clinical and environmental risk factors (e.g., childhood disadvantage, family history of SUD, childhood conduct problems, childhood depression, early exposure to substances, frequent use during adolescence) found it useful in identifying those with persistent SUDs ^11^.

More recently, polygenic scores (PGS), which aggregate risk for a trait across the genome using information from genome-wide association studies (GWAS), were robustly associated with substance use ^12^ and substance related problems ^13^ across adolescence and into young adulthood. However, though robustly associated, current PGS do poorly in identifying individuals affected by SUDs ^14^. To date, there is limited work on the combined impact of genetic, environmental, and clinical risk factors for SUDs. Prior work combining individual genetic variants and clinical features outperformed clinical features alone ^15^, but individual variants have limited predictive power. In other medical conditions, such as melanoma ^16^ or ischemic stroke ^17^, combining clinical and genetic risk factors showed improvement predicting risk for a specific outcome over models using individual risk factors.

In the current study, we examine the joint association of early life clinical/environmental risk factors and PGS with SUDs in early adulthood across four longitudinal cohorts: the National Longitudinal Study of Adolescent to Adult Health (Add Health); the Avon Longitudinal Study of Parents and Children (ALSPAC); the Collaborative Study on the Genetics of Alcoholism (COGA); and the youngest cohort of the Finnish Twin Cohort Study (FinnTwin12). These samples include population-based cohorts from three countries (United States, England, and Finland) and a predominantly high-risk sample. Two of the samples (COGA and Add Health) are ancestrally diverse. We focus on early adulthood as this is a critical period for the development and onset of SUDs ^18^. Our research questions are guided by the understanding that risk factors for SUDs range across multiple levels of analysis.

## METHODS

### Samples

*Add Health* is a nationally representative longitudinal study of adolescents followed into adulthood in the United States ^19^. Data have been collected from Wave I when respondents were between 11-18 (1994-1995) to Wave V (2016-2018) when respondents were 35-42. The current analysis uses data from Waves I, II, and Wave IV.

*ALSPAC* is an ongoing, longitudinal population-based study of a birth cohort in the (former) Avon district of Southwest England ^20–23^. Pregnant female residents with an expected date of delivery between April 1, 1991 and December 31, 1992 were invited to participate (N = 14,541 pregnant women, 80% of those eligible). This analysis uses data up to the age 24 assessment (details of all the data that is available through a searchable, web-based tool: http://www.bristol.ac.uk/alspac/researchers/our-data/).

*COGA* is a family-based sample consisting of alcohol dependent individuals (identified through treatment centers across the United States), their extended families, and community controls (N ∼16,000) ^24,25^. We use a prospective sample of offspring of the original COGA participants (baseline ages 12-22, N = 3,573) that have been assessed biennially since recruitment (2004-2019) ^26^.

*FinnTwin12* is a population-based study of Finnish twins born 1983–1987 identified through Finland’s Central Population Registry. A total of 2,705 families (87% of all identified) returned the initial family questionnaire late in the year in which twins reached age 11 ^27^. Twins were invited to participate in follow-up surveys when they were ages 14, 17, and approximately 22.

Each cohort includes a wide range of social, behavioral, and phenotypic data measured across the life course. The SUD measures were derived from the corresponding young adult phases of data collection in each cohort (mean ages ∼ 22 - 28). A full description of each sample is presented in the supplementary information (section 2).

### Measures

#### Lifetime Diagnosis of Substance Use Disorder

We constructed measures of lifetime SUD diagnosis based on the data that were available in each of the samples, defined as meeting criteria for four, non-mutually exclusive categories of substance dependence: 1) alcohol dependence; 2) nicotine dependence; 3) drug dependence (inclusive of drugs such as cannabis, cocaine, opioids, sedatives, etc.); and 4) any substance dependence (alcohol, nicotine, or drug). Our analyses focused primarily on DSM-IV as this diagnostic system was most consistently used across all samples. There was one exception: in each of the samples, nicotine dependence was measured using a cutoff of 7 or higher on the Fagerstrom Test for Nicotine Dependence (FTND) ^28^. Where possible, we drew measures of substance dependence from data collected during young adulthood to try and maintain temporal ordering between SUD diagnoses and measured risk factors.

#### Clinical/Environmental Risk Index

We created a clinical/environmental risk index (CERI) considering a variety of established risk factors for SUD (Table 1). The CERI included ten validated early life risk factors associated with later development of SUDs, including: low childhood socioeconomic status (SES), family history of SUD, early initiation of substance use, childhood internalizing problems, childhood externalizing problems, frequent drinking in adolescence, frequent smoking in adolescence, frequent cannabis use in adolescence, peer substance use, and exposure to trauma/traumatic experiences ^11,29,30^. We dichotomized each risk factor (present vs not present) and summed them into an index for each person ranging from 0 to 10, providing a single measure of aggregate risk. Dichotomizing these items allowed us to harmonize measures across each sample in an interpretable manner. A full list of how each measure is defined within each of the samples is available in the supplementary information (section 3).

**Table 1:** Items included in the Clinical/Environmental Risk Index (CERI)

| **Measure** | **Definition** |
| --- | --- |
| 1) Low childhood SES | Parent(s) report having less than basic level of education [culturally dependent]; having a low-skill or menial occupation; income at or below the poverty line; or receipt of government assistance. |
| 2) Family history of SUD | Biological parent self-reports history of SUD for themselves or other biological parent or meets criteria for SUD from clinical interview/AUDIT threshold of 8 or higher. |
| 3) Childhood externalizing problems | Respondent meets criteria for conduct disorder or oppositional defiant disorder from a clinical interview or computer-based prediction; or has a behavior problems score at or above the 90th percentile at 15 or younger. |
| 4) Childhood internalizing problems | Respondent reports diagnosis of depression/anxiety or panic disorder; meets criteria for internalizing disorder in clinical interview/computer-based prediction; or has a CES-D score above a threshold of 16 at 15 or younger. |
| 5) Early initiation of substance use | Respondent reports age of first whole alcoholic drink, smoked whole cigarette, or tried cannabis before the age of 15. |
| 6) Adolescent alcohol use | Frequency of self-reported use 5 or more days per week at age 18 and below. |
| 7) Adolescent tobacco use | Frequency of self-reported use at daily use at age 18 and below. |
| 8) Adolescent cannabis use | Frequency of self-reported use 5 or more days per week at age 18 and below. |
| 9) Peer substance use | Respondent reports the majority of their best friends use alcohol/tobacco/cannabis; their three best friends smoke daily/drink once a month/use cannabis once a month; or more than one friend smokes/drinks alcohol/has tried other drugs. |
| 10) Traumatic events | Respondent reports exposure to any traumatic event. |

Full description of sample specific definitions available in the supplementary information.

#### Polygenic Scores

We constructed polygenic scores (PGS), which are aggregate measures of the number of risk alleles individuals carry weighted by effect sizes from GWAS summary statistics, from six recent GWAS of SUDs and comorbid conditions including: 1) externalizing problems (EXT) ^31^; 2) major depressive disorder (MDD) ^32^; 3) problematic alcohol use ^33^ (ALCP); 4) alcohol consumption (drinks per week, ALCC) ^34,35^; 5) cigarettes per day/FTND (CPD) ^34,36^; and 6) schizophrenia (SCZ) ^37,38^. We focused on these PGS, specifically, because: 1) SUDs show strong genetic overlap with other externalizing ^39–41^, internalizing ^32,42^, and psychotic disorders ^33,43,44^; 2) both shared and substance-specific genetic risk are associated with later SUDs ^45–47^; and 3) substance use and SUDs have only partial genetic overlap ^48,49^. Therefore, our PGS cover a spectrum of genetic risk for SUDs, using the most current and well-powered results for each of the listed domains (see supplementary information section 4 for a detailed description).

GWAS have been overwhelmingly limited to individuals of European ancestries ^50,51^. Importantly, PGS derived from GWAS of one ancestry do not always transport into other ancestral populations ^52,53^. We therefore used PRS-CSx ^54^, a new method that combines information from well-powered GWAS (typically of European ancestries) and ancestrally matched GWAS to improve the predictive power of PGS in the African ancestry samples from Add Health and COGA. PRS-CSx integrates GWAS summary statistics across multiple input populations and employs a Bayesian approach to correct GWAS summary statistics for the non-independence of SNPs in linkage disequilibrium (LD) with one another^54^. For participants of European ancestries, we used the EUR derived PRS-CSx results, while we used the EUR+AFR meta-analyzed results for the African ancestry participants. See the supplementary information (section 5) for details.

#### Analytic Strategy

We pooled all the data for analysis using a fixed effects integrative data analytic (IDA) approach ^55^. The IDA approach is more powerful than traditional meta-analyses when one has access to raw data for each of the contributing samples. Our approach to harmonization and pooling was as follows. First, we defined the measures and cutoffs to be used in each of the samples, creating the CERI, PGS, and SUD outcomes at the cohort level. Second, within each cohort, we regressed each PGS on age, age^2^, sex, sex*age, sex*age^2^, and the first 10 ancestral PCs (specific to each sample) to account for population stratification in the PGS. Next, we pooled all the data for analysis. We included cohort as a fixed effect for each of the six cohorts (4 samples, of which two were split by ancestry) in subsequent analyses. Additionally, we included age of last observation and sex as covariates.

We estimated a series of nested logistic regression models with the pooled data: 1) a baseline model (sex, age, and cohort), 2) a genetic risk model (baseline + PGS), 3) a clinical/environmental risk model (baseline + CERI), and 4) a combined risk model (baseline + PGS + CERI). Because COGA and FT12 included a large number of related individuals, we adjusted for familial clustering using cluster-robust standard errors ^56^. To assess the predictive accuracy of each model, we took the difference in pseudo-*R*^*2*^ (Δ*Pseudo-R*^*2*^) ^57^, between the baseline and corresponding models. Finally, we calculated the discriminatory power of the combined model using the area under the curve (AUC) from a receiver operating characteristic (ROC) curve. We included a variety of robustness checks to ensure that no single cohort in the IDA was unduly influencing the results. Our analytic strategy was preregistered on the Open Science Framework (https://osf.io/etbw8). Deviations from the preregistration are described in the supplementary information (section 6).

## RESULTS

Table 2 contains the descriptive statistics for each of the cohorts and ancestries. Each cohort had similar proportions of females (∼51% - 56%). The mean ages ranged from ∼22 to ∼29 years of age. The COGA cohorts (both European and African ancestries) reported the highest rates of SUD, an expected finding given the nature of the sample (highly selected for SUDs). Add Health participants generally had higher rates of SUD than ALSPAC or FinnTwin12, but lower than COGA. Finally, ALSPAC and FinnTwin12 reported similar levels of alcohol, nicotine, drug, and any substance dependence. COGA participants reported higher mean values on the CERI. The remaining cohorts report relatively similar rates of risk factor exposure.

**Table 2:**
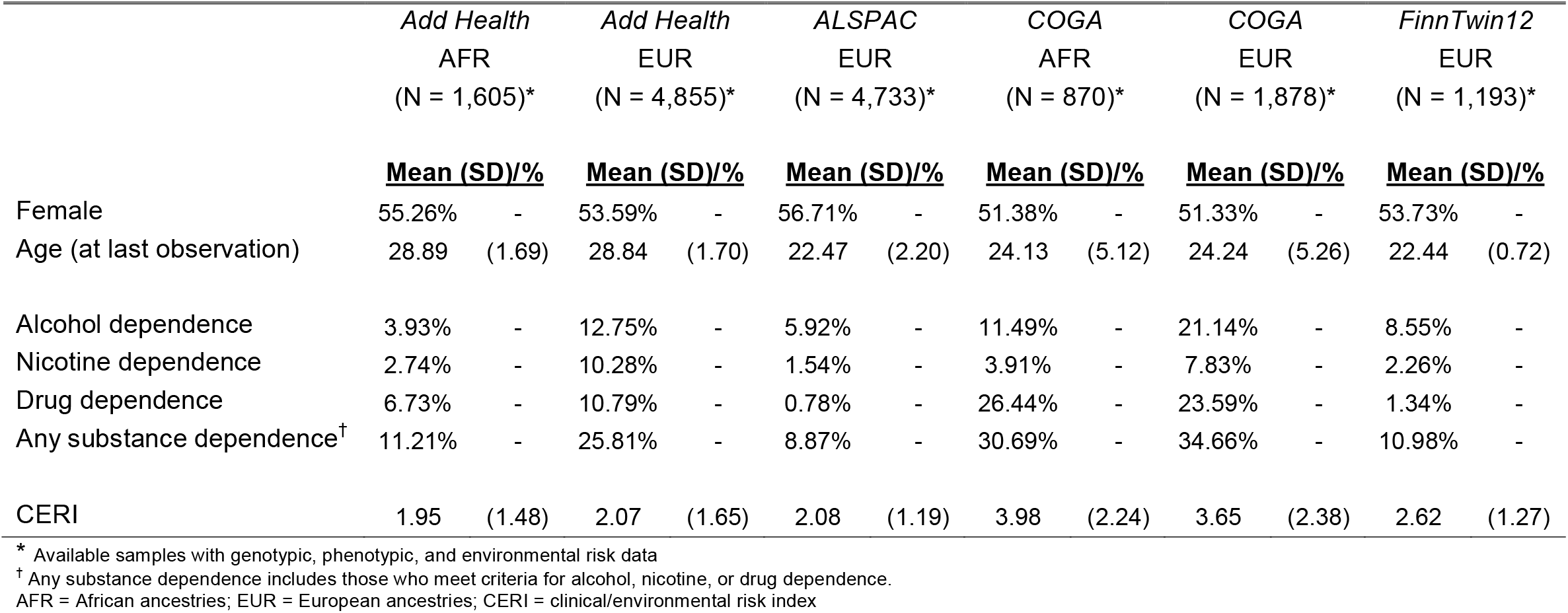
Prevalence of SUDs and CERI by Cohort.

|  | *Add Health*<br>AFR<br>(N = 1,605)* |  | *Add Health*<br>EUR<br>(N = 4,855)* |  | *ALSPAC*<br>EUR<br>(N = 4,733)* |  | *COGA*<br>AFR<br>(N = 870)* |  | *COGA*<br>EUR<br>(N = 1,878)* |  | *FinnTwin12*<br>EUR<br>(N = 1,193)* |  |
| --- | --- | --- | --- | --- | --- | --- | --- | --- | --- | --- | --- | --- |
|  | <u>Mean (SD)/%</u> |  | <u>Mean (SD)/%</u> |  | <u>Mean (SD)/%</u> |  | <u>Mean (SD)/%</u> |  | <u>Mean (SD)/%</u> |  | <u>Mean (SD)/%</u> |  |
| Female | 55.26% | - | 53.59% | - | 56.71% | - | 51.38% | - | 51.33% | - | 53.73% | - |
| Age (at last observation) | 28.89 | (1.69) | 28.84 | (1.70) | 22.47 | (2.20) | 24.13 | (5.12) | 24.24 | (5.26) | 22.44 | (0.72) |
| Alcohol dependence | 3.93% | - | 12.75% | - | 5.92% | - | 11.49% | - | 21.14% | - | 8.55% | - |
| Nicotine dependence | 2.74% | - | 10.28% | - | 1.54% | - | 3.91% | - | 7.83% | - | 2.26% | - |
| Drug dependence | 6.73% | - | 10.79% | - | 0.78% | - | 26.44% | - | 23.59% | - | 1.34% | - |
| Any substance dependence<sup>†</sup> | 11.21% | - | 25.81% | - | 8.87% | - | 30.69% | - | 34.66% | - | 10.98% | - |
| CERI | 1.95 | (1.48) | 2.07 | (1.65) | 2.08 | (1.19) | 3.98 | (2.24) | 3.65 | (2.38) | 2.62 | (1.27) |

\* Available samples with genotypic, phenotypic, and environmental risk data
<sup>†</sup> Any substance dependence includes those who meet criteria for alcohol, nicotine, or drug dependence.
AFR = African ancestries; EUR = European ancestries; CERI = clinical/environmental risk index

Table 3 presents the results from the *PGS only, CERI only*, and *combined* models for each outcome. Three of the six PGS were associated with the SUD outcomes in the *PGS only* model. EXT was associated with each of the SUD outcomes (EXT OR = 1.18 – 1.50); ALCP was associated with alcohol dependence and any substance dependence (ALCP OR = 1.10 – 1.13); and CPD was associated with nicotine dependence (CPD OR = 1.33). In the *CERI only* models, the CERI was consistently associated across each of the SUD categories (ORs = 1.37 – 1.67). When we combined the PGS and CERI into the same model, the CERI remained significant across SUDs and was largely unchanged (ORs = 1.35 – 1.65). EXT remained associated with drug dependence (OR = 1.11) and nicotine dependence (OR = 1.33), ALCP remained associated alcohol dependence (OR = 1.12), and CPD remained associated with nicotine dependence (OR = 1.31). Both EXT and ALCP remained associated with any substance dependence diagnosis (ORs = 1.09 – 1.18). Overall, the combined model explained 5.9%, 12.6%, 13.1%, and 12.8% of the variance in alcohol dependence, nicotine dependence, drug dependence, and any substance dependence, respectively.

**Table 3:** Estimates for PGS Only, CERI Only, and Combined Models.

|  |  | Alcohol<br>Dependence |  |  | Nicotine<br>Dependence |  |  | Drug<br>Dependence |  |  | Any substance<br>dependence |  |  |
| --- | --- | --- | --- | --- | --- | --- | --- | --- | --- | --- | --- | --- | --- |
|  |  | <u>OR</u> | <u>95%</u> | <u>CI</u> | <u>OR</u> | <u>95%</u> | <u>CI</u> | <u>OR</u> | <u>95%</u> | <u>CI</u> | <u>OR</u> | <u>95%</u> | <u>CI</u> |
| PGS Only Model* | ALCC PGS | 1.05 | (0.99, | 1.11) | 0.96 | (0.89, | 1.04) | 1.05 | (0.98, | 1.12) | 1.00 | (0.96, | 1.05) |
|  | ALCP PGS | **1.13** | **(1.06,** | **1.20)** | 1.01 | (0.93, | 1.10) | 1.07 | (1.00, | 1.15) | **1.10** | **(1.05,** | **1.16)** |
|  | EXT PGS | **1.18** | **(1.11,** | **1.26)** | **1.50** | **(1.38,** | **1.63)** | **1.27** | **(1.19,** | **1.36)** | **1.31** | **(1.25,** | **1.38)** |
|  | MDD PGS | 1.00 | (0.94, | 1.06) | 1.06 | (0.98, | 1.15) | 1.08 | (1.02, | 1.15) | 1.02 | (0.98, | 1.07) |
|  | SCZ PGS | 1.04 | (0.97, | 1.10) | 0.98 | (0.90, | 1.06) | 1.03 | (0.96, | 1.11) | 1.00 | (0.96, | 1.05) |
|  | CPD PGS | 1.00 | (0.94, | 1.06) | **1.33** | **(1.24,** | **1.43)** | 1.01 | (0.95, | 1.08) | 1.08 | (1.03, | 1.13) |
| | $\Delta Pseudo-R^2$ | | 0.011 | | | 0.037 | | | 0.014 | | | 0.022 | |
| CERI Only Model* | CERI | **1.37** | **(1.33,** | **1.41)** | **1.63** | **(1.57,** | **1.70)** | **1.67** | **(1.61,** | **1.72)** | **1.58** | **(1.54,** | **1.63)** |
| | $\Delta Pseudo-R^2$ | | 0.054 | | | 0.107 | | | 0.129 | | | 0.120 | |
| Combined Model* | CERI | **1.35** | **(1.31,** | **1.40)** | **1.58** | **(1.52,** | **1.65)** | **1.65** | **(1.59,** | **1.70)** | **1.55** | **(1.51,** | **1.60)** |
|  | ALCC PGS | 1.04 | (0.97, | 1.10) | 0.94 | (0.87, | 1.03) | 1.03 | (0.96, | 1.11) | 0.99 | (0.94, | 1.04) |
|  | ALCP PGS | **1.12** | **(1.05,** | **1.19)** | 0.99 | (0.91, | 1.08) | 1.06 | (0.98, | 1.14) | **1.09** | **(1.04,** | **1.15)** |
|  | EXT PGS | 1.08 | (1.01, | 1.15) | **1.33** | **(1.22,** | **1.45)** | **1.11** | **(1.03,** | **1.20)** | **1.18** | **(1.12,** | **1.24)** |
|  | MDD PGS | 0.97 | (0.91, | 1.03) | 1.02 | (0.94, | 1.10) | 1.03 | (0.96, | 1.10) | 0.98 | (0.93, | 1.03) |
|  | SCZ PGS | 1.03 | (0.97, | 1.10) | 0.96 | (0.88, | 1.05) | 1.01 | (0.94, | 1.08) | 1.00 | (0.95, | 1.05) |
|  | CPD PGS | 0.98 | (0.92, | 1.04) | **1.31** | **(1.22,** | **1.42)** | 0.98 | (0.92, | 1.04) | 1.06 | (1.01, | 1.11) |
| | $\Delta Pseudo-R^2$ | | 0.059 | | | 0.126 | | | 0.131 | | | 0.128 | |

\* All models included age, sex, and cohort as covariates. See Supplementary Table 7 for all parameter estimates. PGS residualized on age, sex, and first 10 ancestral principal components.
Bolded estimates = $p < .05$ after correction for multiple testing ( $p < .05/4 = 0.0125$ )
$\Delta Pseudo-R^2$ denotes pseudo- $R^2$ above model including age, sex, and cohort. CI = confidence interval; PGS = polygenic score; CERI = clinical/environmental risk index

Figure 1 (Panel A) presents the raw prevalence for each outcome across counts of the CERI. The proportion of those meeting criteria for SUDs among those reporting 3 or more, 5 or more, and 7 or more risk factors surpassed lifetime prevalence estimates from nationally representative samples for drug dependence, alcohol dependence, and nicotine dependence, respectively ^58^. Panel B depicts the prevalence of each category of SUD across several mutually exclusive categories: 1) those in the bottom 90% of both the CERI and all PGS (averaged across the six scores); 2) those in the top 10% of the CERI but the bottom 90% of the PGS distribution; 3) those in the top 10% of the PGS distribution and the bottom 90% of the CERI; and 4) those in the top 10% of both PGS and the CERI. There is an increase in risk across those with elevated genetic risk, clinical/environmental risk, and both. Those in the top 10% of both PGS and CERI had the highest prevalence of each of the SUDs, though the error bars overlap with the estimates from those in the top 10% of the risk index, alone. Compared to those in the bottom 90% on both, those in the to the top 10% of both have a relative risk of 3.86 (95% CI = 3.20, 4.65) for alcohol dependence, 6.11 (95% CI = 4.84, 7.72) for nicotine dependence, 8.04 (95% CI = 6.92, 9.36) for drug dependence, and 4.05 (95% CI = 3.64, 4.51) for any substance dependence.

**Figure 1:**
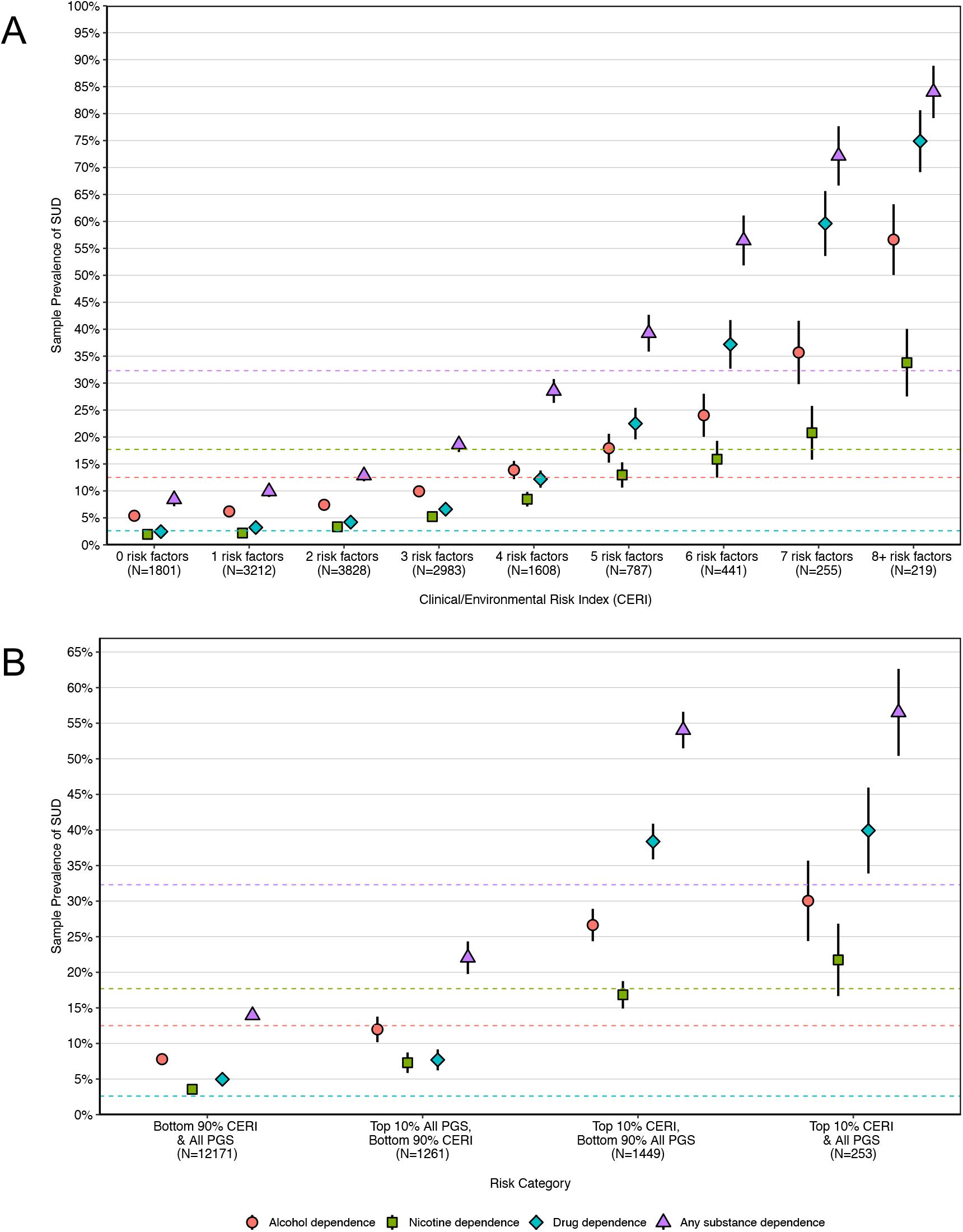
SUD Prevalence Across Genetic and Environmental Risk Factors. Panel A: Prevalence (and 95% confidence intervals) of those who meet criteria for alcohol, nicotine, drug, or any substance dependence across counts for items in the risk index. Panel B: Prevalence (and 95% confidence intervals) of those who meet criteria for alcohol, nicotine, drug, or any substance dependence across four categories: 1) those below the 90^th^ percentile for all PGS and the CERI; 2) those at or above the 90^th^ percentile for the CERI; 3) those at or above the 90^th^ percentile for all PGS; and 4) those at or above the 90^th^ percentile for both the CERI and PGS. PGS and risk index were first residualized on sex, age, age^2^, cohort, sex*age, sex*age^2^, sex*cohort, cohort*age, cohort*age^2^, sex*cohort*age, and sex*cohort*age^2^. Dotted colored lines represent corresponding lifetime prevalence estimates for alcohol dependence (red), nicotine dependence (green), drug dependence (blue), and any substance use disorder (purple) from nationally representative data ^58^.

Finally, we considered the AUC for the combined model for each of the SUD categories. Figure 2 presents the ROC curves for the full (CERI and PGS) and baseline (covariates only) models for each SUD category. The AUC for each combined model was 0.74 for alcohol dependence, 0.82 for nicotine dependence, 0.86 for drug dependence, and 0.78 for any substance dependence. The overall change in AUC (from the baseline to the full model) that we achieve when adding the CERI and PGS was modest (ΔAUC = 0.05 – 0.10), and this improvement was due in large part to the explanatory power of the CERI. ROC curves for the CERI only and PGS only models are presented in Supplemental Figure 6.

**Figure 2:**
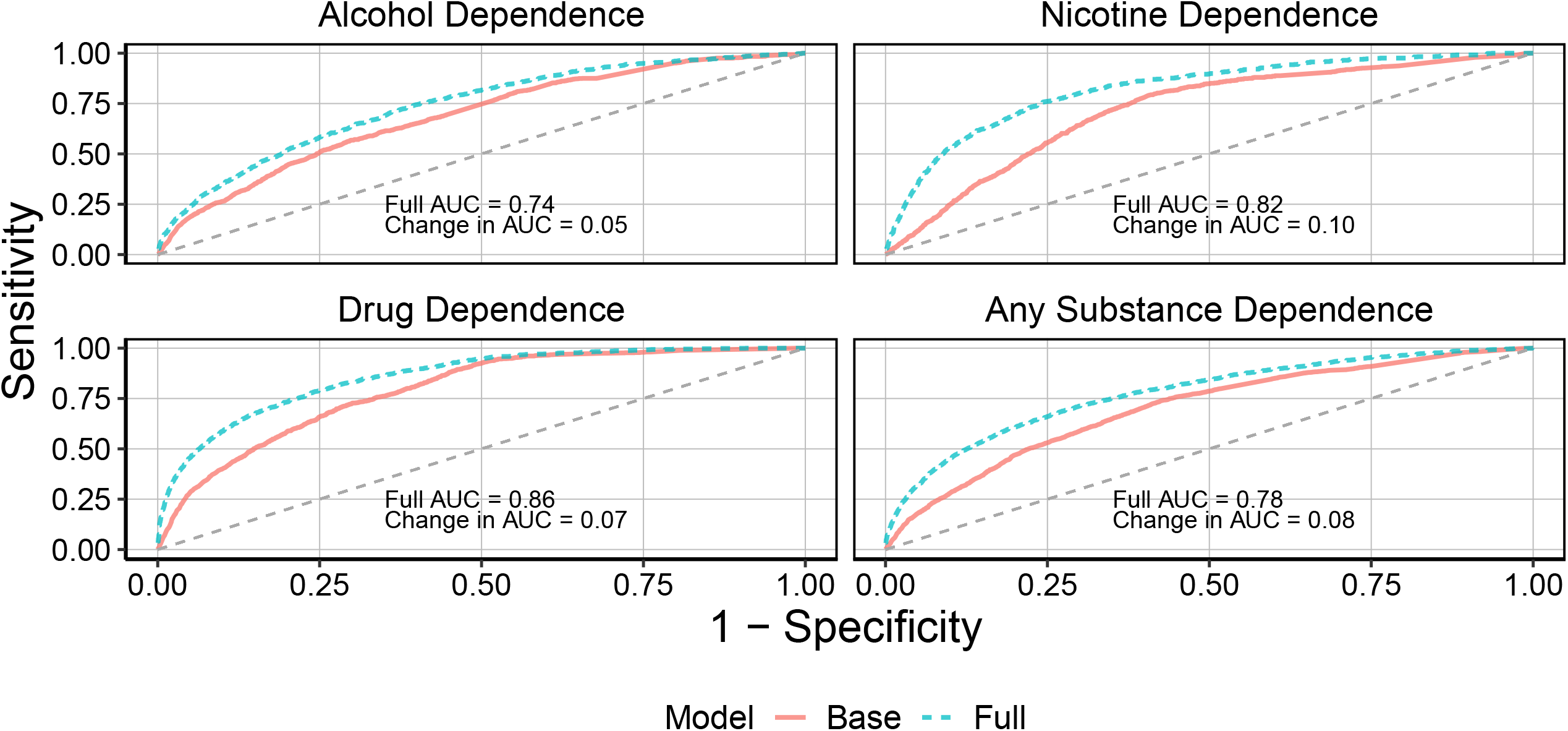
ROC Curves for Combined and Baseline Models. Receiver operating characteristic (ROC) curves for baseline models (red line, covariates only) and the full models (blue line, PGS + CERI + covariates) for each substance use disorder. Area under the curve (AUC) is presented for the PGS model in each cell. Change in AUC represents value of the difference between AUC from the full model and AUC from the base model.

### Sensitivity Analyses

We performed a variety of sensitivity analyses. Results from leave-one-out (LOO) and sex-stratified analyses were largely similar to those from the main results. In ancestry stratified analyses, results in the cohorts of European ancestries largely mirrored the main results. None of the PGS were associated with SUDs in the cohorts of African ancestries. Effect sizes for the CERI were largely similar across European and African ancestries (see Supplemental Tables S1-S3) and were mostly stable when removing individual risk factors (supplemental information section 7).

We also tested for interactions between the PGS and CERI and cohort (Add Health EUR as the reference group). There were few significant interactions and no consistent patterns in variation for PGS, though the CERI did show considerable variation across cohort (Supplemental Table S4). Finally, we fit complimentary models using a random effects approach, allowing the slopes for the PGS and CERI to vary randomly across cohort. Random slopes for PGS did not consistently improve model fit, though a random slope for the CERI consistently improved model fit (Supplemental Table S5). We compared the parameter estimates from the random effect models to the main analyses and results were largely consistent (Supplemental Table S6).

## DISCUSSION

Substance use disorders remain a serious threat to public health ^59^. In the current analysis, we examined the combination of clinical, environmental, and genetic risk factors for determining who is more likely to develop a SUD in early adulthood. We used previously validated measures of environmental and clinical risk ^11,29,30^ and polygenic scores for externalizing problems ^31^, major depressive disorder ^32^, problematic alcohol use ^33,35^, alcohol consumption ^34,35^, cigarettes per day/nicotine dependence ^34,36^, and schizophrenia ^37,38^. The combination of genetic and social-environmental measures was significantly associated with the development of SUDs. The overall association was strongest for drug dependence, followed by any substance dependence, nicotine dependence, and alcohol dependence.

The CERI was the strongest association with each outcome. The proportion of those meeting criteria for each SUD surpassed lifetime estimates in persons with 3 or more, 5 or more, and 7 or more risk factors for drug dependence, alcohol dependence, and nicotine dependence, respectively. The discriminatory power of the combined model (AUC = .74 - .86) was similar to AUC estimates published in the original paper from which many of the risk index items were derived (AUC ∼ 0.80) ^11^. Interestingly, this risk index was originally developed for identifying persons with persistent SUD through early mid-life (∼age 40). In the current analysis we demonstrated that the CERI in conjunction with demographic covariates and PGS does equally well for those who meet criteria for any SUD by young adulthood.

The overall predictive power of the PGS alone was in the range of 1.1 – 3.7%. Only the PGS for externalizing problems, problematic alcohol use, and cigarettes per day were consistently associated with SUD outcomes. The PGS for externalizing problems was associated with drug dependence and nicotine dependence, the PGS for problematic alcohol use PGS was associated with alcohol dependence, and both were associated with any substance dependence. The PGS for cigarettes per day was only associated with nicotine dependence. Overall, these results support prior evidence that genetic risk for SUDs consists of a both shared and substance-specific variance ^31,41,47^.

Interestingly, even though the effect sizes were attenuated in the model, the PGS for externalizing problems, problematic alcohol use, and cigarettes per day remained significantly associated when we included the CERI, though the additional information the PGS provided was minimal. Since the CERI also included many of the phenotypes each of the PGS measured (e.g., childhood conduct disorder for externalizing, childhood depression for major depressive disorder; and frequent alcohol use for alcohol consumption), part of this attenuation is likely due to the inclusion of the actual phenotypes through which risk for some of these disorders is expressed. PGS are also confounded by environmental variance ^59^ and the reduction in effect sizes could be accounting for some of that confounding. PGS may add information beyond well-known risk factors, which could prove useful when information on certain exposures or behaviors is unavailable.

Further refinement of risk measures may improve our ability to develop screening protocols for those at greater risk of developing substance-related problems. Early detection has the potential to improve prevention efforts, as prior work suggests that those at highest risk of substance misuse stand to benefit the most from prevention efforts ^60^. Ideally, screening tools for SUD risk would include measures of social, clinical, and genetic risk factors, as each impacts the development of SUDs ^5–7,61,62^. In the push for precision medicine, very often the focus is on biological information, but social determinants of health are also critically important.

Currently, these tools are not ready for clinical use. If we reach the point where social, clinical, and genetic information become sufficiently powerful, we must recognize that identifying persons for early intervention carries a significant risk. Screening for social determinants has the potential for unintended consequences, including further stigmatization ^63^. Genetic information has even more potential for abuse and stigmatization. Policy makers must ensure that there is comprehensive legal protection against discrimination using any form of information. Additionally, any attempt to use social, clinical, or genetic information for targeted intervention or identification in a clinical setting must be done so in a patient-centered approach, rather than any “one-size fits all” that exclude patients from their own healthcare decisions ^64^.

Our analysis has several important limitations. First, although we included individuals of diverse ancestries, the PGS for our samples of African ancestries were severely underpowered due to the small size of the discovery sample. Large-scale GWAS in diverse cohorts are vital to ensuring that any benefit of precision medicine is shared equitably across the population ^65^. Second, while distinct, ancestry is related to race-ethnicity, and with it, racism and racial discrimination, some of the most profound social determinants of health ^66^. Our measure of environmental risk was crude and may not fully capture risk factors that contribute to SUDs in populations beyond non-Hispanic Whites. Future studies should include racially relevant measures of risk (e.g., experiences of interpersonal racism/discrimination, racial residential segregation) as well as other social and environmental measures that are known risk factors for SUDs (e.g., neighborhood social conditions, alcohol outlet density). Further refinement of known risk factors may allow for better prediction of those at risk of developing an SUD. We did observe variation in the predictive ability of the CERI across cohorts, suggesting the observed effect may differ in magnitude across populations. We therefore urge caution in overinterpreting study results. Finally, while we tried to ensure time order between risk factors and onset of disorder, some risk factors (particularly adolescent substance use) could have occurred concurrently with diagnosis. Future work in samples with risk factors measured before the initiation of substance use (such as the Adolescent Brain Cognitive Development Study) will be important for replication efforts.

Recognizing that multiple social, clinical, and genetic factors contribute to risk for SUDs is important as we move towards the goal precision medicine that benefits all segments of the population. There is still much work to be done before tools such as these are useful in a clinical setting. However, the results of this integrative data analysis provide initial evidence *all* of these risk factors contribute unique information to SUDs in early adulthood. Expanding our sources of information (such as electronic health records, census data from home of record) and making use of increasingly well-powered PGS will continue to improve our ability to understand how SUDs develop.

## Data Availability

All data in the present study are available upon application and approval by the constituent cohorts.

https://osf.io/fg9rj/

https://osf.io/etbw8

## ACKNOWLEDGEMENTS

Research reported in this publication was supported by the National Institute on Alcohol Abuse and Alcoholism and the National Institute of Drug Abuse of the National Institutes of Health under award numbers R01AA015416, R01DA050721, R01DA042090, and K02AA018755; the Academy of Finland (grants 100499, 205585, 118555, 141054, 265240, 308248, 308698 and 312073); and the Scientific and Technological Research Council of Turkey (TÜBİTAK) under award number 114C117 (FA); and the Sigrid Juselius Foundation. The content is solely the responsibility of the authors and does not necessarily represent the official views of any of the funding bodies. This research also used summary data from the Psychiatric Genomics Consortium (PGC), the Million Veterans Program (MVP), the GWAS and Sequencing Consortium for Alcohol and Nicotine (GSCAN), UK Biobank, the Genomic Psychiatry Cohort (GPC) and 23andMe, Inc. We would like to thank the many studies that made these consortia possible, the researchers involved, and the participants in those studies, without whom this effort would not be possible. We would also like to thank the research participants and employees of 23andMe.

**The Externalizing Consortium:** Principal Investigators: Danielle M. Dick, Philipp Koellinger, K. Paige Harden, Abraham A. Palmer. Lead Analysts: Richard Karlsson Linnér, Travis T. Mallard, Peter B. Barr, Sandra Sanchez-Roige. Significant Contributors: Irwin D. Waldman. The Externalizing Consortium has been supported by the National Institute on Alcohol Abuse and Alcoholism (R01AA015416 - administrative supplement), and the National Institute on Drug Abuse (R01DA050721). Additional funding for investigator effort has been provided by K02AA018755, U10AA008401, P50AA022537, as well as a European Research Council Consolidator Grant (647648 EdGe to Koellinger). The content is solely the responsibility of the authors and does not necessarily represent the official views of the above funding bodies. **Add Health:** Add Health is directed by Robert A. Hummer and funded by the National Institute on Aging cooperative agreements U01 AG071448 (Hummer) and U01AG071450 (Aiello and Hummer) at the University of North Carolina at Chapel Hill. Waves I-V data are from the Add Health Program Project, grant P01 HD31921 (Harris) from *Eunice Kennedy Shriver* National Institute of Child Health and Human Development (NICHD), with cooperative funding from 23 other federal agencies and foundations. Add Health was designed by J. Richard Udry, Peter S. Bearman, and Kathleen Mullan Harris at the University of North Carolina at Chapel Hill. **ALSPAC:** We are extremely grateful to all the families who took part in this study, the midwives for their help in recruiting them, and the whole ALSPAC team, which includes interviewers, computer and laboratory technicians, clerical workers, research scientists, volunteers, managers, receptionists, and nurses. The UK Medical Research Council and Wellcome (Grant ref: 217065/Z/19/Z) and the University of Bristol provide core support for ALSPAC. This publication is the work of the authors, and Peter Barr and Danielle Dick will serve as guarantors for the contents of this paper. A comprehensive list of grants funding is available on the ALSPAC website (http://www.bristol.ac.uk/alspac/external/documents/grant-acknowledgements.pdf); This research was specifically funded by the Medical Research Council (MRC) under grants MR/L022206/1, MR/M006727/1, and G0800612/86812; the Wellcome Trust under grant 086684; and the National Institute on Alcohol Abuse and Alcoholism under 5R01AA018333-05. GWAS data was generated by Sample Logistics and Genotyping Facilities at Wellcome Sanger Institute and LabCorp (Laboratory Corporation of America) using support from 23andMe. **COGA:** We thank The Collaborative Study on the Genetics of Alcoholism (COGA), Principal Investigators B. Porjesz, V. Hesselbrock, T. Foroud; Scientific Director, A. Agrawal; Translational Director, D. Dick, includes eleven different centers: University of Connecticut (V. Hesselbrock); Indiana University (H.J. Edenberg, T. Foroud, Y. Liu, M.H. Plawecki); University of Iowa Carver College of Medicine (S. Kuperman, J. Kramer); SUNY Downstate Health Sciences University (B. Porjesz, J. Meyers, C. Kamarajan, A. Pandey); Washington University in St. Louis (L. Bierut, J. Rice, K. Bucholz, A. Agrawal); University of California at San Diego (M. Schuckit); Rutgers University (J. Tischfield, R. Hart, J. Salvatore); The Children’s Hospital of Philadelphia, University of Pennsylvania (L. Almasy); Virginia Commonwealth University (D. Dick); Icahn School of Medicine at Mount Sinai (A. Goate, P. Slesinger); and Howard University (D. Scott). Other COGA collaborators include: L. Bauer (University of Connecticut); J. Nurnberger Jr., L. Wetherill, X., Xuei, D. Lai, S. O’Connor, (Indiana University); G. Chan (University of Iowa; University of Connecticut); D.B. Chorlian, J. Zhang, P. Barr, S. Kinreich, G. Pandey (SUNY Downstate); N. Mullins (Icahn School of Medicine at Mount Sinai); A. Anokhin, S. Hartz, E. Johnson, V. McCutcheon, S. Saccone (Washington University); J. Moore, Z. Pang, S. Kuo (Rutgers University); A. Merikangas (The Children’s Hospital of Philadelphia and University of Pennsylvania); F. Aliev (Virginia Commonwealth University); H. Chin and A. Parsian are the NIAAA Staff Collaborators. We continue to be inspired by our memories of Henri Begleiter and Theodore Reich, founding PI and Co-PI of COGA, and also owe a debt of gratitude to other past organizers of COGA, including Ting-Kai Li, P. Michael Conneally, Raymond Crowe, and Wendy Reich, for their critical contributions. This national collaborative study is supported by NIH Grant U10AA008401 from the National Institute on Alcohol Abuse and Alcoholism (NIAAA) and the National Institute on Drug Abuse (NIDA). All code necessary to replicate this study is available upon request.

## ETHICS DECLARATIONS

The authors have no conflicts of interest to declare.

## DATA AVAILABILITY

All data sources are described in the manuscript and supplemental information. No new data were collected. Only data from existing studies or study cohorts were analyzed, some of which have restricted access to protect the privacy of the study participants. Add Health genetic data obtained through dbGaP (Study Accession: phs001367.v1.p1). Instructions on gaining access to Add Health restricted use data can be found at: https://data.cpc.unc.edu/projects/2/view. COGA genetic data available through dbGaP (Study Accession: phs000763.v1.p1). Instructions for access to ALSPAC data available at: http://www.bristol.ac.uk/alspac/researchers/access/. The process for obtaining the GWAS summary statistics used in these analyses are described in the corresponding original GWAS publications.

## CODE AVAILABILITY

No custom algorithms or software was developed in this study. All code is available by request from the corresponding author. Polygenic scores generated using PRS-CSx (https://github.com/getian107/PRScsx). All primary analyses completed in R 4.1.0 using the *data*.*table* (1.14.0), *pROC* (1.18.0), *lme4* (1.1-27.1), and base packages.

## REFERENCES

1. U.S. Overdose Deaths In 2021 Increased Half as Much as in 2020 - But Are Still Up 15%. https://www.cdc.gov/nchs/pressroom/nchs_press_releases/2022/202205.htm.

2. Degenhardt, L. et al. The global burden of disease attributable to alcohol and drug use in 195 countries and territories, 1990–2016: a systematic analysis for the Global Burden of Disease Study 2016. The Lancet Psychiatry 5, 987–1012 (2018).

3. Sacks, J. J., Gonzales, K. R., Bouchery, E. E., Tomedi, L. E. & Brewer, R. D. 2010 National and State Costs of Excessive Alcohol Consumption. American Journal of Preventive Medicine 49, e73–e79 (2015).

4. National Drug Intelligence Center. National drug threat assessment. vol. 2019 (2011).

5. Verhulst, B., Neale, M. C. & Kendler, K. S. The heritability of alcohol use disorders: a meta-analysis of twin and adoption studies. Psychol Med 45, 1061–1072 (2015).

6. Verweij, K. J. H. et al. Genetic and environmental influences on cannabis use initiation and problematic use: A meta-analysis of twin studies. Addiction 105, 417–430 (2010).

7. Kendler, K. S., Jacobson, K. C., Prescott, C. A. & Neale, M. C. Specificity of Genetic and Environmental Risk Factors for Use and Abuse/Dependence of Cannabis, Cocaine, Hallucinogens, Sedatives, Stimulants, and Opiates in Male Twins. American Journal of Psychiatry 160, 687–695 (2003).

8. Galea, S., Nandi, A. & Vlahov, D. The Social Epidemiology of Substance Use. Epidemiologic Reviews 26, 36–52 (2004).

9. Barr, P. B. Neighborhood conditions and trajectories of alcohol use and misuse across the early life course. Health and Place 51, 36–44 (2018).

10. Barr, P. B., Silberg, J., Dick, D. M. & Maes, H. H. Childhood socioeconomic status and longitudinal patterns of alcohol problems: Variation across etiological pathways in genetic risk. Social Science and Medicine 209, 51–58 (2018).

11. Meier, M. H. et al. Which adolescents develop persistent substance dependence in adulthood? Using population-representative longitudinal data to inform universal risk assessment. Psychol Med 46, 877–889 (2016).

12. Schaefer, J. D. et al. Associations between polygenic risk of substance use and use disorder and alcohol, cannabis, and nicotine use in adolescence and young adulthood in a longitudinal twin study. Psychological Medicine 1–11 (2021) doi:10.1017/S0033291721004116.

13. Deak, J. D. et al. Alcohol and nicotine polygenic scores are associated with the development of alcohol and nicotine use problems from adolescence to young adulthood. Addiction 117, 1117–1127 (2022).

14. Barr, P. B. et al. Using polygenic scores for identifying individuals at increased risk of substance use disorders in clinical and population samples. Translational Psychiatry 10, 196 (2020).

15. Kinreich, S. et al. Predicting risk for Alcohol Use Disorder using longitudinal data with multimodal biomarkers and family history: a machine learning study. Molecular Psychiatry 26, 1133–1141 (2021).

16. Gu, F. et al. Combining common genetic variants and non-genetic risk factors to predict risk of cutaneous melanoma. Human Molecular Genetics (2018) doi:10.1093/hmg/ddy282.

17. O’Sullivan, J. W. et al. Combining clinical and polygenic risk improves stroke prediction among individuals with atrial fibrillation. medRxiv 2020.06.17.20134163 (2020) doi:10.1101/2020.06.17.20134163.

18. Kessler, R. C. et al. Lifetime prevalence and age-of-onset distributions of DSM-IV disorders in the National Comorbidity Survey Replication. Archives of General Psychiatry 62, 593 (2005).

19. Harris, K. M., Halpern, C. T., Haberstick, B. C. & Smolen, A. The National Longitudinal Study of Adolescent Health (Add Health) sibling pairs data. Twin Research and Human Genetics 16, 391–398 (2013).

20. Boyd, A. et al. Cohort profile: The ‘Children of the 90s’-The index offspring of the avon longitudinal study of parents and children. International Journal of Epidemiology (2013) doi:10.1093/ije/dys064.

21. Fraser, A. et al. Cohort profile: The avon longitudinal study of parents and children: ALSPAC mothers cohort. International Journal of Epidemiology 42, 97–110 (2013).

22. Harris, P. A. et al. Research electronic data capture (REDCap)-A metadata-driven methodology and workflow process for providing translational research informatics support. Journal of Biomedical Informatics 42, 377–381 (2009).

23. Northstone, K. et al. The Avon Longitudinal Study of Parents and Children (ALSPAC): an update on the enrolled sample of index children in 2019 [version 1; peer review: 2 approved]. Wellcome Open Research 4, (2019).

24. Edenberg, H. J. The collaborative study on the genetics of alcoholism: An update. Alcohol Research and Health 26, 214–218 (2002).

25. Begleiter, H. The Collaborative Study on the Genetics of Alcoholism. Alcohol Health and Research World 19, 228 (1995).

26. Bucholz, K. K. et al. Comparison of Parent, Peer, Psychiatric, and Cannabis Use Influences Across Stages of Offspring Alcohol Involvement: Evidence from the COGA Prospective Study. Alcoholism: Clinical and Experimental Research 41, 359–368 (2017).

27. Rose, R. J. R. J. et al. FinnTwin12 Cohort: An Updated Review. Twin Research and Human Genetics vol. 22 (2019).

28. Heatherton, T. F., Kozlowski, L. T., Frecker, R. C. & Fagerstrom, K. O. The Fagerstrom Test for Nicotine Dependence: a revision of the Fagerstrom Tolerance Questionnaire. Br J Addict 86, 1119–1127 (1991).

29. Hughes, K. et al. The effect of multiple adverse childhood experiences on health: a systematic review and meta-analysis. The Lancet Public Health (2017) doi:10.1016/S2468-2667(17)30118-4.

30. Sher, K. J., Grekin, E. R. & Williams, N. A. The development of alcohol use disorders. Annual Review of Clinical Psychology 1, 493–523 (2005).

31. Karlsson Linner, R. et al. Multivariate genomic analysis of 1.5 million people identifies genes related to addiction, antisocial behavior, and health. Nature Neuroscience.

32. Levey, D. F. et al. Bi-ancestral depression GWAS in the Million Veteran Program and meta-analysis in >1.2 million individuals highlight new therapeutic directions. Nature Neuroscience (2021) doi:10.1038/s41593-021-00860-2.

33. Zhou, H. et al. Genome-wide meta-analysis of problematic alcohol use in 435,563 individuals yields insights into biology and relationships with other traits. Nature Neuroscience (2020) doi:10.1038/s41593-020-0643-5.

34. Liu, M. et al. Association studies of up to 1.2 million individuals yield new insights into the genetic etiology of tobacco and alcohol use. Nature Genetics 51, 237–244 (2019).

35. Kranzler, H. R. et al. Genome-wide association study of alcohol consumption and use disorder in 274,424 individuals from multiple populations. Nature Communications 10, 1499 (2019).

36. Quach, B. C. et al. Expanding the genetic architecture of nicotine dependence and its shared genetics with multiple traits. Nature Communications 11, (2020).

37. Trubetskoy, V. et al. Mapping genomic loci implicates genes and synaptic biology in schizophrenia. Nature 2022 1–13 (2022) doi:10.1038/s41586-022-04434-5.

38. Bigdeli, T. B. et al. Genome-Wide Association Studies of Schizophrenia and Bipolar Disorder in a Diverse Cohort of US Veterans. Schizophrenia Bulletin (2020) doi:10.1093/schbul/sbaa133.

39. Barr, P. B. & Dick, D. M. The Genetics of Externalizing Problems. Current Topics in Behavioral Neurosciences 47, 93–112 (2020).

40. Krueger, R. F. et al. Etiological connections among substance dependence, antisocial behavior and personality: Modeling the externalizing spectrum. J Abnorm Psychol 111, 411–424 (2002).

41. Kendler, K. S. & Myers, J. The boundaries of the internalizing and externalizing genetic spectra in men and women. Psychological Medicine 44, 647–655 (2014).

42. Polimanti, R. et al. Evidence of causal effect of major depression on alcohol dependence: Findings from the psychiatric genomics consortium. Psychological Medicine (2019) doi:10.1017/S0033291719000667.

43. Johnson, E. C. et al. A large-scale genome-wide association study meta-analysis of cannabis use disorder. The Lancet Psychiatry (2020) doi:10.1016/S2215-0366(20)30339-4.

44. Zhou, H. et al. Association of OPRM1 Functional Coding Variant With Opioid Use Disorder: A Genome-Wide Association Study. JAMA Psychiatry (2020) doi:10.1001/jamapsychiatry.2020.1206.

45. Kendler, K. S., Gardner, C. & Dick, D. M. Predicting alcohol consumption in adolescence from alcohol-specific and general externalizing genetic risk factors, key environmental exposures and their interaction. Psychol Med 41, 1507–1516 (2011).

46. Meyers, J. L. et al. Genetic Influences on Alcohol Use Behaviors Have Diverging Developmental TrajectoriesL: A Prospective Study Among Male and Female Twins. Alcoholism: Clinical and Experimental Research 38, 2869–2877 (2014).

47. Barr, P. B. et al. Parsing Genetically Influenced Risk Pathways: Genetic Loci Impact Problematic Alcohol Use Via Externalizing and Specific Risk. medRxiv 2021.07.20.21260861 (2021) doi:10.1101/2021.07.20.21260861.

48. Sanchez-Roige, S., Palmer, A. A. & Clarke, T. K. Recent Efforts to Dissect the Genetic Basis of Alcohol Use and Abuse. Biological Psychiatry (2020) doi:10.1016/j.biopsych.2019.09.011.

49. Walters, R. K. et al. Trans-ancestral GWAS of alcohol dependence reveals common genetic underpinnings with psychiatric disorders. Nature Neuroscience 21, 1656–1669 (2018).

50. Dick, D. M., Barr, P., Guy, M., Nasim, A. & Scott, D. Review: Genetic research on alcohol use outcomes in African American populations: A review of the literature, associated challenges, and implications. American Journal on Addictions vol. 26 486–493 (2017).

51. Mills, M. C. & Rahal, C. A scientometric review of genome-wide association studies. Communications Biology 2, 9 (2019).

52. Martin, A. R. et al. Human Demographic History Impacts Genetic Risk Prediction across Diverse Populations. Am J Hum Genet 100, 635–649 (2017).

53. Duncan, L. et al. Analysis of polygenic risk score usage and performance in diverse human populations. Nature Communications (2019) doi:10.1038/s41467-019-11112-0.

54. Ruan, Y. et al. Improving Polygenic Prediction in Ancestrally Diverse Populations. Nature Genetics (2022) doi:10.1038/s41588-022-01054-7.

55. Curran, P. J. & Hussong, A. M. Integrative Data Analysis: The Simultaneous Analysis of Multiple Data Sets. Psychological Methods 14, 81–100 (2009).

56. Cameron, C. A., Gelbach, J. B. & Miller, D. L. Robust inference with multiway clustering. Journal of Business and Economic Statistics 29, 238–249 (2011).

57. Nagelkerke, N. J. D. A note on a general definition of the coefficient of determination. Biometrika 78, 691–692 (1991).

58. Hasin, D. S. & Grant, B. F. The National Epidemiologic Survey on Alcohol and Related Conditions (NESARC) Waves 1 and 2: review and summary of findings. Social Psychiatry and Psychiatric Epidemiology 50, 1609–1640 (2015).

59. Conrod, P. J. et al. Effectiveness of a Selective, Personality-Targeted Prevention Program for Adolescent Alcohol Use and Misuse: A Cluster Randomized Controlled Trial. JAMA Psychiatry 70, 334–342 (2013).

60. Kong, A. et al. The nature of nurture: Effects of parental genotypes. Science (1979) 359, 424–428 (2018).

61. Burt, S. A. Are there meaningful etiological differences within antisocial behavior? Results of a meta-analysis. Clin Psychol Rev 29, 163–178 (2009).

62. Rhee, S. H. & Waldman, I. D. Genetic and environmental influences on antisocial behavior: A meta-analysis of twin and adoption studies. Psychol Bull 128, 490–529 (2002).

63. Garg, A., Boynton-Jarrett, R. & Dworkin, P. H. Avoiding the Unintended Consequences of Screening for Social Determinants of Health. JAMA 316, 813–814 (2016).

64. Davidson, K. W. & McGinn, T. Screening for Social Determinants of Health: The Known and Unknown. JAMA 322, 1037–1038 (2019).

65. Martin, A. R. et al. Clinical use of current polygenic risk scores may exacerbate health disparities. Nature Genetics 51, 584–591 (2019).

66. Williams, D. R., Mohammed, S. A., Leavell, J. & Collins, C. Race, socioeconomic status, and health: complexities, ongoing challenges, and research opportunities. Ann N Y Acad Sci 1186, 69–101 (2010).

