## Supplemental information for "Clinical, Environmental, and Genetic Risk Factors for Substance Use Disorders: Characterizing Combined Effects across Multiple Cohorts"

### Table of contents

*1 Study introduction ... 3*

*2 Samples ... 4*

**2.1 The National Longitudinal Study of Adolescent to Adult Health (Add Health) ... 4**

**2.2 Avon Longitudinal Study of Parents and Children (ALSPAC) ... 4**

**2.3 The Collaborative Study of the Genetics of Alcoholism (COGA) ... 5**

**2.4 The Finnish Twin Cohort (FinnTwin12) ... 5**

*3 Clinical/environmental risk index measures ... 7*

**3.1 Low childhood socioeconomic status (SES) ... 7**

**3.2 Family history of substance use disorders (SUD) ... 8**

**3.3 Childhood behavior/externalizing problems ... 9**

**3.4 Childhood internalizing problems ... 9**

**3.5 Early substance use initiation ... 10**

**3.6 Frequent adolescent alcohol use ... 10**

**3.7 Frequent adolescent tobacco use ... 11**

**3.8 Frequent adolescent cannabis use ... 12**

**3.9 Adolescent peer substance use ... 12**

**3.10 Exposure to stressful/traumatic events ... 13**

*4 GWAS selection and inclusion ... 17*

**4.1 GWAS of externalizing (EXT) ... 17**

**4.2 GWAS of major depressive disorder (MDD) ... 17**

**4.3 GWAS of problematic alcohol use (ALCP) ... 17**

**4.4 GWAS of alcohol consumption (ALCC) ... 18**

**4.5 GWAS of schizophrenia (SCZ) ... 18**

**4.6 GWAS of cigarettes per day/nicotine dependence (CPD) ... 18**

*5 Polygenic Score Creation ... 19*

*6 Deviations from preregistration ... 21*

*7 Variation in effect of clinical/environmental risk index (CERI) ... 22*

*8 ROC Curves for CERI only and PGS only Models ... 24*

*9 Random-effects Integrative Data Analysis (RE IDA) ... 26*

*10 References ... 27*

### 1 Study introduction

Substance use disorders (SUD) are associated with substantial cost to society, affected individuals, and their families. In 2016, alcohol use contributed 4.2% to the global disease burden and other drug use contributed 1.3%[1]. Excessive alcohol use is estimated to cost the United States $250 billion[2] every year. Illicit drugs cost the United States approximately $190 billion[3] annually, of which $78.5 billion is due to opioid misuse, alone[4]. Given the substantial human and economic costs of misuse and disorders, developing methods of identifying persons at heightened risk for SUD is a vital public health concern.

Ideally, screening tools for SUD risk would include measures of environmental, clinical, and genetic risk factors, as each are known to impact the development of substance use disorders [5–9]. Previous research using an index of established clinical and environmental risk factors related to adult SUD (e.g., childhood disadvantage, family history of SUD, childhood conduct problems, childhood depression, early exposure to substances, frequent use during adolescence) found this risk index to be useful (AUC ~ .80) in differentiating between individuals that were affected and unaffected with SUDs[10]. For measures of genetic risk, recent analyses evaluating the potential for polygenic risk scores, or PGS, which aggregate risk for a trait across the genome using information from genome-wide association studies (GWAS), have found current PGS alone provide little additional information to differentiate between individuals affected and unaffected by SUDs[11]. However, no research has examined these genetic, environmental, and clinical risk factors for SUD together. For other medical conditions, such as melanoma[12] or ischemic stroke[13], models using combined clinical and genetic risk factors showed improvement over models using individual risk factors in isolation.

The current proposal builds upon prior work developing risk indices for SUDs. We examined the joint effect of early life (defined as the periods of childhood and adolescence) risk factors and genetic liability (in the form of polygenic risk scores) to build prediction models for lifetime diagnosis of SUDs (alcohol dependence, drug dependence, and/or any substance dependence) using four longitudinal cohorts: the Collaborative Study on the Genetics of Alcoholism (COGA); the National Longitudinal Study of Adolescent to Adult Health (Add Health); the Avon Longitudinal Study of Parents and Children (ALSPAC); and the younger cohort of the Finnish Twin Study (FinnTwin12; FT12). We performed all analyses according to a preregistered analysis plan, which was time-stamped on December 3, 2020 (https://osf.io/etbw8).

### 2 Samples

#### 2.1 The National Longitudinal Study of Adolescent to Adult Health (Add Health)

*Add Health* is an ongoing, nationally representative longitudinal study of adolescents followed into adulthood in the United States[14]. Data has been collected ranging from Wave I when respondents were between 11-18 (1994-1995) to Wave V (2016-2018) when respondents were 35-42. Add Health participants were selected from a stratified sample of 132 schools resulting in an initial, nationally representative sample of 90,118 students in grades 7-12. Of the original sample, 20,745 were selected for additional in-home interviews. Of those who completed the Wave I interview (1994-1995), 14,738 (71%) completed Wave II (1996); 15,197 (73%) completed Wave III (2001-2002); and 15,701 (75%) completed Wave IV (2007-2008). Most respondents completed the majority of the waves, with 16,278 (78%) completing three or more waves. Wave V (ages 32-42) data collection is underway, with a target sample of 19,828 (data for N = 3,872 is already released). In total, 15,159 individuals interviewed during Wave IV (ages 24-32) provided samples for genotyping, conducted using the Illumina Omni1 and Omni2.5 arrays. After quality control, genotypic data are available for 9,974 individuals (5,896 non-Hispanic White; 2,081 African American; 1,448 Hispanic; 550 Other). Genotypes for European ancestry participants were imputed to the Haplotype Reference Consortium (HRC) reference panel[15], and data for the African ancestry were imputed to the 1000 Genomes, Phase III reference panel[16]. The current analysis uses data from Waves I and II, when respondents were adolescents, and Wave IV, when respondents received a clinical interview assessing lifetime SUD diagnosis. We removed those who were >18 years old at Wave I to ensure timing of childhood/adolescent risk factors. Our final analytic sample consisted of 4,855 individuals of European ancestries and 1,605 individuals of African ancestries.

#### 2.2 Avon Longitudinal Study of Parents and Children (ALSPAC)

*ALSPAC* is an ongoing, longitudinal population-based study of a birth cohort in the (former) Avon district of Southwest England[17–19]. Study data were collected and managed using REDCap electronic data capture tools hosted at the University of Bristol. REDCap (Research Electronic Data Capture) is a secure, web-based software platform designed to support data capture for research studies [20]. Pregnant women resident in Avon, UK with expected dates of delivery 1st April 1991 to 31st December 1992 were invited to take part in the study. The initial number of pregnancies enrolled is 14,541 (for these at least one questionnaire has been returned or a “Children in Focus” clinic had been attended by 19/07/99). Of these initial pregnancies, there was a total of 14,676 fetuses, resulting in 14,062 live births and 13,988 children who were alive at 1 year of age.

When the oldest children were approximately 7 years of age, an attempt was made to bolster the initial sample with eligible cases who had failed to join the study originally. As a result, when considering variables collected from the age of seven onwards (and potentially abstracted from obstetric notes) there are data available for more than the 14,541 pregnancies mentioned above. The number of new pregnancies not in the initial sample (known as Phase I enrollment) that are currently represented on the built files and reflecting enrollment status at the age of 24 is 913 (456, 262 and 195 recruited during Phases II, III and IV respectively), resulting in an additional 913 children being enrolled. The phases of enrollment are described in more detail in the cohort profile paper and its update (see footnote 4 below). The total sample size for analyses using any data collected after the age of seven is therefore 15,454 pregnancies, resulting in 15,589 fetuses. Of these 14,901 were alive at 1 year of age.

A 10% sample of the ALSPAC cohort, known as the Children in Focus (CiF) group, attended clinics at the University of Bristol at various time intervals between 4 to 61 months of age. The CiF group were chosen at random from the last 6 months of ALSPAC births (1432 families attended at least one clinic). Excluded were those mothers who had moved out of the area or were lost to follow-up, and those partaking in another study of infant development in Avon.

Ethical approval for the study was obtained from the ALSPAC Ethics and Law Committee and the Local Research Ethics Committee. Consent for biological samples has been collected in accordance with the Human Tissue Act (2004). Informed consent for the use of data collected via questionnaires and clinics was obtained from participants following the recommendations of the ALSPAC Ethics and Law Committee at the time. Children from the ALSPAC cohort were genotyped using the Illumina HumanHap550 quad chip genotyping platform[21]. Genotype data were imputed to the Haplotype Reference Consortium (HRC) reference panel[15]. Our final analytic sample consisted of 4,733 individuals of European ancestries.

#### 2.3 The Collaborative Study of the Genetics of Alcoholism (COGA)

COGA, initiated in 1989 to identify genes associated with vulnerability for AUD, ascertained *high-risk families* through adult probands in treatment for alcohol dependence[22]. Probands along with all willing first-degree relatives were assessed; recruitment was extended to include additional relatives in families that contained 2 or more first degree relatives with alcohol dependence and community- ascertained comparison families (n = 16,848). Data collection included a psychiatric interview (the Semi-Structured Assessment for the Genetics of Alcoholism, or SSAGA[23]), neurophysiological and neuropsychological protocols, and collection of blood for DNA. We currently have genome wide data on 12,145 individuals (8,038 individuals of European ancestry; 3,655 individuals of African ancestry). In 2004, COGA began the prospective study of adolescents and young adults, targeting assessment of youth aged 12-22 from COGA families where at least one parent had been interviewed[24]. These subjects were re-assessed every two years; currently, 89% of individuals have 2+ interviews. COGA is *racially/ethnically diverse* (60.6% non-Hispanic White, 24.9% African American, 11.1% Hispanic, and 3.4% Other). Genotyping of the COGA samples was conducted across different phases of data collection. European ancestry (EA) samples were genotyped at multiple sites, including: (1) Center for Inherited Disease Research using the Illumina HumanHap1M array; (2) Genome Technology Access Center at Washington University School of Medicine using the Illumina OmniExpress; and (3) Rutgers University using the Affymetrix Smokescreen array. In addition, the two datasets genotyped on the Smokescreen genotyping array were also imputed separately, due to different processing pipelines used by the genotyping laboratory. Principal components were computed from GWAS data using Eigenstrat and 1000 Genomes, Phase III reference panel[16]. Individual ancestry was assigned using the YRI, CEU, JPT and CHB populations to set reference points. We limited our focus to the prospective sample of adolescent and young adult offspring (bassline ages 12-22; N = 3,573) of the original phases of COGA adult participants in the current analyses. Our final analytic sample consisted of 1,878 individuals of European ancestries and 870 individuals of African ancestries.

#### 2.4 The Finnish Twin Cohort (FinnTwin12)

*FinnTwin12* is the youngest cohort of the Finnish Twin Cohort Study, a population-based study of Finnish twins born 1983–1987 identified through Finland's Central Population Registry. A total of 2,705 families (87% of all identified) returned the initial family questionnaire late in the year in which twins reached age 11[25]. Twins were invited to participate in follow-up surveys when they were ages 14, 17, and approximately 22 (during young adulthood). An intensively studies sample was selected as 1035 families, among whom 1854 twins were interviewed at age 14. The interviewed twins were invited as young adults to complete the Semi-Structured Assessment for the Genetics of Alcoholism (SSAGA)[23] interview (n = 1,347) and provide DNA samples[26]. Genotyping was conducted using the Human670-QuadCustom Illumina BeadChip at the Wellcome Trust Sanger Institute. Quality control steps included removing SNPs with minor allele frequency (MAF) <1%, genotyping success rate <95%, or Hardy–Weinberg equilibrium $p < 1 \times 10^{-6}$, and removing individuals with genotyping success rate <95%, a mismatch between phenotypic and genotypic gender, excess relatedness (outside of known families), and heterozygosity outliers. Genotypes were imputed to the Haplotype Reference Consortium (HRC) reference panel[15]. The current analysis uses data from the intensive sub sample with available DNA and diagnostic data across each wave of data collection. Our final analytic sample consisted of 1,193 individuals of European ancestries.

### 3 Clinical/environmental risk index measures

The environmental/clinical risk index was based on a previously validated index of risk factors for persistent SUD[10], including low childhood socioeconomic status (SES), family history of SUD, early initiation of substance use, childhood internalizing problems, childhood externalizing problems, frequent drinking in adolescence, frequent smoking in adolescence, frequent cannabis use in adolescence, along with other known risk factors, such as peer substance use[54], and exposure to trauma/traumatic experiences[55]. We dichotomized each risk factor (present vs not present) and summed them into an index for each person ranging from 0 to 10, providing a single measure of aggregate risk. In order to ensure that constructs were comparable across each of the four samples, we compared and harmonized the available measures. Below, we present the exact measurement for each of the ten items in each sample. Supplemental Figure 2 depicts the breakdown of each risk factor across each of the cohorts. Supplemental Figure 3 presents the tetrachoric correlations between each of the risk factors, by cohort and pooled into one sample. While there is variation in the strength of the correlations, overwhelmingly we see that many of these risk factors are weakly-to-modestly, positively correlated with one another. The strongest correlations (~.7) are between frequent tobacco and cannabis use in adolescence. Even this relatively strong correlation suggests that, at most, ~50% of the variance is shared between any given item in the risk index. The lack of consistent, strong correlations indicate that these items are not mere proxies for one another.

#### 3.1 Low childhood socioeconomic status (SES)

##### *3.1.1 Add Health*

Participants were classified as experiencing low SES in childhood if they met criteria for any of the below items:

(i) Parental education: both residential parents reported having less than a high school.

(ii) Parental occupation: both residential parents reported occupations that were manual/low wage/low skill.

(iii) Household poverty: respondents report household income at or below the 1994 Federal Poverty threshold (Poverty Status: 1 person/Per extra person/4 person HH example = 7360/2480/14800).

(iv) Receipt of public assistance: respondent or parents report receipt of public assistance.

##### *3.1.2 ALSPAC*

Participants were classified as experiencing low SES in childhood if they met criteria for any of the below items:

(i) Parental education: mother and partner (if present) report no educational qualifications.

(ii) Household poverty: mother reported weekly income less than 100 pounds a week at ages 2.5, 4, or 7.

###### *3.1.3 COGA*

Participants were classified as experiencing low SES in childhood if their parent(s) reported having less than a high school.

###### *3.1.4 FinnTwin12*

Participants were classified as experiencing low SES in childhood if they met criteria for any of the below items:

(i) Parental education: parent(s) reported having less than a basic level education (minimum in Finland).

(ii) Parental occupation: both parents reported occupations that were manual/low wage/low skill.

#### 3.2 Family history of substance use disorders (SUD)

###### *3.2.1 Add Health*

Respondents were classified as having a family history of SUD if parents reported yes to either of the following questions:

(i) "Does {NAME]'s biological mother currently have the following health problem (check all that apply): Alcoholism"

(ii) "Does {NAME]'s biological father currently have the following health problem (check all that apply): Alcoholism"

###### *3.2.2 ALSPAC*

Respondents were classified as having a family history of SUD if parents met criteria for any of the below items:

(i) Mother/Father - AUDIT total score greater than a threshold of 8.

(ii) Mother/Father - Self-reported having alcoholism or a drug addiction.

###### *3.2.3 COGA*

Respondents were classified as having a family history of SUD if parents met criteria for an alcohol use disorder based on parent SSAGA interviews. In instances where direct parent SSAGA interview is not available, collateral parental alcohol use disorder information collected as part of family history reports was used[24,56].

###### *3.2.4 FinnTwin12*

Respondents were classified as having a family history of SUD if parents met criteria for any substance use disorder based on parent SSAGA interviews.

#### 3.3 Childhood behavior/externalizing problems

##### *3.3.1 Add Health*

Respondents were classified as having childhood behavior problems if their score on a list of antisocial behaviors was at or above the $90^{th}$ percentile.

##### *3.3.2 ALSPAC*

Respondents were classified as having childhood behavior problems if participants met DSM-IV clinical diagnostic criteria for any oppositional-conduct disorder.

##### *3.3.3 COGA*

Respondents were classified as having childhood behavior problems if they met criteria for conduct disorder (CD) or oppositional defiant disorder (ODD) from the SSAGA/C-SSAGA interview.

##### *3.3.4 FinnTwin12*

Respondents were classified as having childhood behavior problems if they met criteria for conduct disorder (CD) or oppositional defiant disorder (ODD) from the age 14 SSAGA interview.

#### 3.4 Childhood internalizing problems

##### *3.4.1 Add Health*

Respondents were classified as having childhood internalizing problems if their score on the Center for Epidemiological Study Depression Scale (CES-D) was above 16 before age 15 or they retrospectively reported a diagnosis of depression from before age 15 at Wave IV.

##### *3.4.2 ALSPAC*

Respondents were classified as having childhood internalizing problems based on the Short Mood and Feelings Questionnaire (SMFQ) scores and Strengths and Difficulties Questionnaire (SDQ) emotional symptoms scores.

##### *3.4.3 COGA*

Respondents were classified as having childhood internalizing problems if they reported an onset age below age 15 on the following item across the SSAGA/C-SSAGA interview:

> (i) "Think about the time in your life that stands out as the "worst" time in your life of feeling (MOOD ENDORSED ABOVE). I'm interested in periods that lasted at least two weeks."

##### *3.4.4 FinnTwin12*

Respondents were classified as having childhood internalizing problems if they met criteria for major depressive disorder (MDD) from the age 14 SSAGA interview.

#### 3.5 Early substance use initiation

##### *3.5.1 Add Health*

Respondents were classified as having initiated substance use early if they reported an age below 15 for any of the following Wave I items, or reported use in the Wave II follow up and their age was below 15:

(i) "How old were you when you smoked a whole cigarette for the first time?"

(ii) "Think about the first time you had a drink of beer, wine, or liquor... How old were you then?"

(iii) "How old were you when you tried marijuana for the first time?"

##### *3.5.2 ALSPAC*

Respondents were classified as having initiated substance use early if they reported an age below 15 for any of the following items across the ages 12.5, 13.5, 15.5, 17.5, or 24 follow-ups:

(i) Age of respondent when first smoked a cigarette

(ii) Age when respondent had first whole alcoholic drink

(iii) Age of respondent when first tried cannabis

##### *3.5.3 COGA*

Respondents were classified as having initiated substance use early if they reported an age below 15 for any of the following items across from the SSAGA/C-SSAGA interviews:

(i) "How old were you the first time you had your very first whole drink?"

(ii) "How old were you the first time you smoked a full cigarette?"

(iii) "How old were you the first time you used marijuana?"

##### *3.5.4 FinnTwin12*

Respondents were classified as having initiated substance use early if they reported an age below 15 for any of the following items across the ages 12, 14, and 17.5 interviews:

(i) Age of respondent when first smoked a cigarette

(ii) Age when respondent had first whole alcoholic drink

(iii) Age of respondent when first tried cannabis.

#### 3.6 Frequent adolescent alcohol use

##### *3.6.1 Add Health*

Respondents were classified as regular users if they reported drinking on most days ($\geq$ 5 days a week) before age 18 (Waves I and II), using the following question:

(i) "During the past 12 months, on how many days did you drink alcohol?"

##### *3.6.2 ALSPAC*

Respondents were classified as regular users if they reported drinking on most days (≥ 5 days a week) before age 18, using the following question:

(i) "How often do you have a drink containing alcohol?"

##### *3.6.3 COGA*

Respondents were classified as regular users if they reported drinking on most days (≥ 5 days a week) before age 18, using any of the following questions:

(i) “On how many days did you drink any beverages containing alcohol during the last 12 months?” (from C-SSAGA interview)

(ii) If respondents reported an onset age before age 18 on the following SSAGA question: “Was there ever a time when you drank almost every day for a week or more?”

##### *3.6.4 FinnTwin12*

Respondents were classified as regular users if they reported drinking on most days (≥ 5 days a week) before age 18 (age 14 and 17 survey), using the following question:

(i) “How often do you drink any amount of alcohol?”

#### 3.7 Frequent adolescent tobacco use

##### *3.7.1 Add Health*

Respondents were classified as regular users if they reported smoking daily before age 18 (Waves I and II), using the following question:

(i) “During the past 30 days, on how many days did you smoke cigarettes?”

##### *3.7.2 ALSPAC*

Respondents were classified as regular users if they reported smoking daily before age 18, using the following questions:

(i) "Please mark the box next to the statement which describes you the best: - I usually smoke one or more cigarettes every day” "

(i) "Do you smoke every day?"

##### *3.7.3 COGA*

Respondents were classified as regular users if they reported smoking daily before age 18, using the following question:

(i) “When were you smoking regularly, how many days per week did you usually smoke cigarettes?”

###### *3.7.4 FinnTwin12*

Respondents were classified as regular users if they reported smoking daily before age 18 (age 14 and 17 survey), using the following question:

(i) "Which of the following best describes your present smoking habits: I smoke at least once each day"

#### 3.8 Frequent adolescent cannabis use

###### *3.8.1 Add Health*

Respondents were classified as regular users if they reported cannabis use on most days (≥ 5 days a week) before age 18 (Waves I and II), using the following question:

(i) "During the past 30 days, how many times did you use marijuana?"

###### *3.8.2 ALSPAC*

Respondents were classified as regular users if they reported cannabis use on most days (≥ 5 days a week) before age 18, using the following questions:

(i) Frequency respondent uses or takes cannabis (example response option "I sometimes use or take cannabis but less than once a week"),

(ii) "How many times per week? (over the last 6 months)"

###### *3.8.3 COGA*

Respondents were classified as regular users if they reported regular use before age 18, using the following question from SSAGA/C-SSAGA:

(i) "How old were you the (first/last) time you used marijuana almost every day for at least two weeks?

###### *3.8.4 FinnTwin12*

Respondents were classified as regular users if they reported cannabis use on most days (≥ 5 days a week) before age 18 from the cannabis section of the age 22 SSAGA (retrospective).

#### 3.9 Adolescent peer substance use

###### *3.9.1 Add Health*

Respondents were classified as having substance using peers if they reported 3 or more of their best friends used substances from the following questions at Waves I and II"

(i) "Of your three best friends, how many smoke at least 1 cigarette a day?"

(ii) "Of your three best friends, how many drink alcohol at least once a month?"

(iii) "Of your three best friends, how many use marijuana at least once a month?"

##### *3.9.2 ALSPAC*

Respondents were classified as having substance using peers if they reported most or all of their friends' used substances from the following items:

(i) Number of friends that drank alcohol during the last year

(ii) Number of friends that smoked cigarettes during the last year

(iii) Number of friends that took illegal drugs during the last year

##### *3.9.3 COGA*

Respondents were classified as having substance using peers if they reported most of their friends' used substances from the following SSAGA/C-SSAGA questions (ages 12 – 17):

(i) C-SSAGA: "How many of your best friends smoke?"; "How many of your best friends use alcohol?"; "How many of your best friends use marijuana?"; and "How many of your best friends use other drugs (like cocaine, uppers, or any of the other drugs we've talked about)?"

(ii) SSAGA (retrospective reports): "When you were 12-17, how many of your best friends smoked?"; "how many of your best friends used alcohol?"; "how many of your best friends used marijuana?"; and "how many of your best friends used other drugs (like cocaine, uppers, or any of the other drugs we've talked about)?"

##### *3.9.4 FinnTwin12*

Respondents were classified as having substance using peers if they reported most of their friends' used substances from the following questions at ages 14 and 17:

(i) "Do any of your friends smoke?"

(ii) "Do any of your friends drink?"

(iii) "Have any of your acquaintances tried drugs?"

#### 3.10 Exposure to stressful/traumatic events

##### *3.10.1 Add Health*

Respondents were classified as having been exposed to a stressful/traumatic event if they reported any of the following:

(i) Friend or family member committed suicide

(ii) Victim of a violent assault, sexual assault (females only), or other violent crime

(iii) Witness violence

(iv) Serious injury

(v) Experience intimate partner violence

(vi) Loss of a child

(vii) Loss of a parent

##### *3.10.2 ALSPAC*

Respondents were classified as having been exposed to a stressful/traumatic event if they reported any of the following:

(ii) ever been physically or sexually abused as a child

(iii) ever been bullied

(iv) ever had a serious illness, injury, or hospitalization

(i) ever experienced the death of a parent, sibling, close friend

##### *3.10.3 COGA*

Respondents were classified as having been exposed to a stressful/traumatic event if they reported any of the following:

(i) Ever been shot

(ii) Ever been stabbed

(iii) Ever been mugged or threatened with a weapon or experienced a break-in or robbery

(iv) Ever been raped or sexually assaulted by a relative

(v) Ever been raped or sexually assaulted by someone not related to you

(vi) Ever been in military combat

(vii) Ever wounded in combat

(viii) Ever been held captive, tortured, or kidnapped

(ix) Ever been in a natural disaster like a fire, flood, earthquake, tornado, mudslide, or hurricane

(x) Ever been in a serious accident

(xi) Ever seen someone being seriously injured or killed

(xii) Ever unexpectedly discovered a dead body

##### *3.10.4 FinnTwin12*

FinnTwin12 did not contain measures related to stressful or traumatic events.

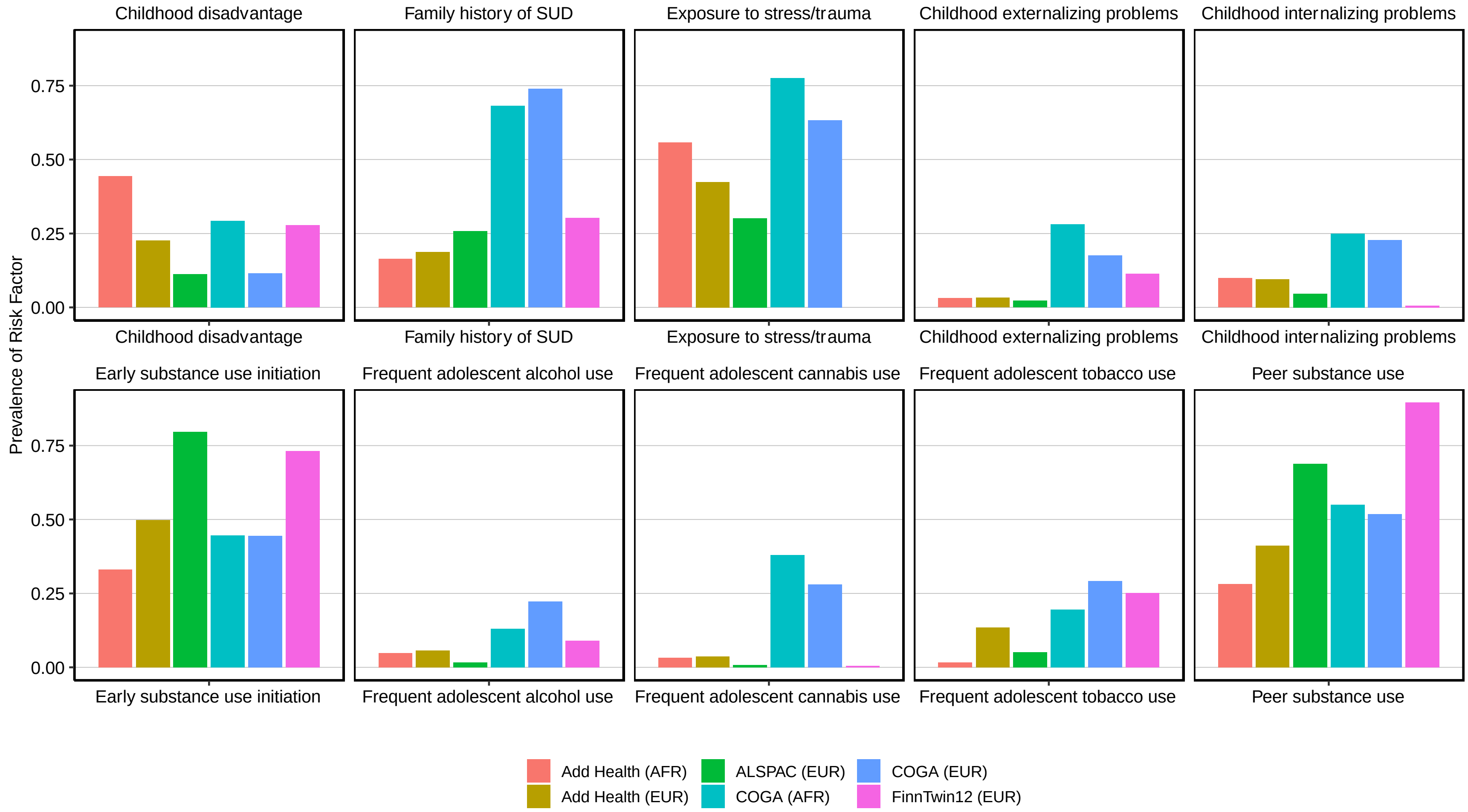


*Supplemental Figure 1: Prevalence of Risk Factors by Cohort*

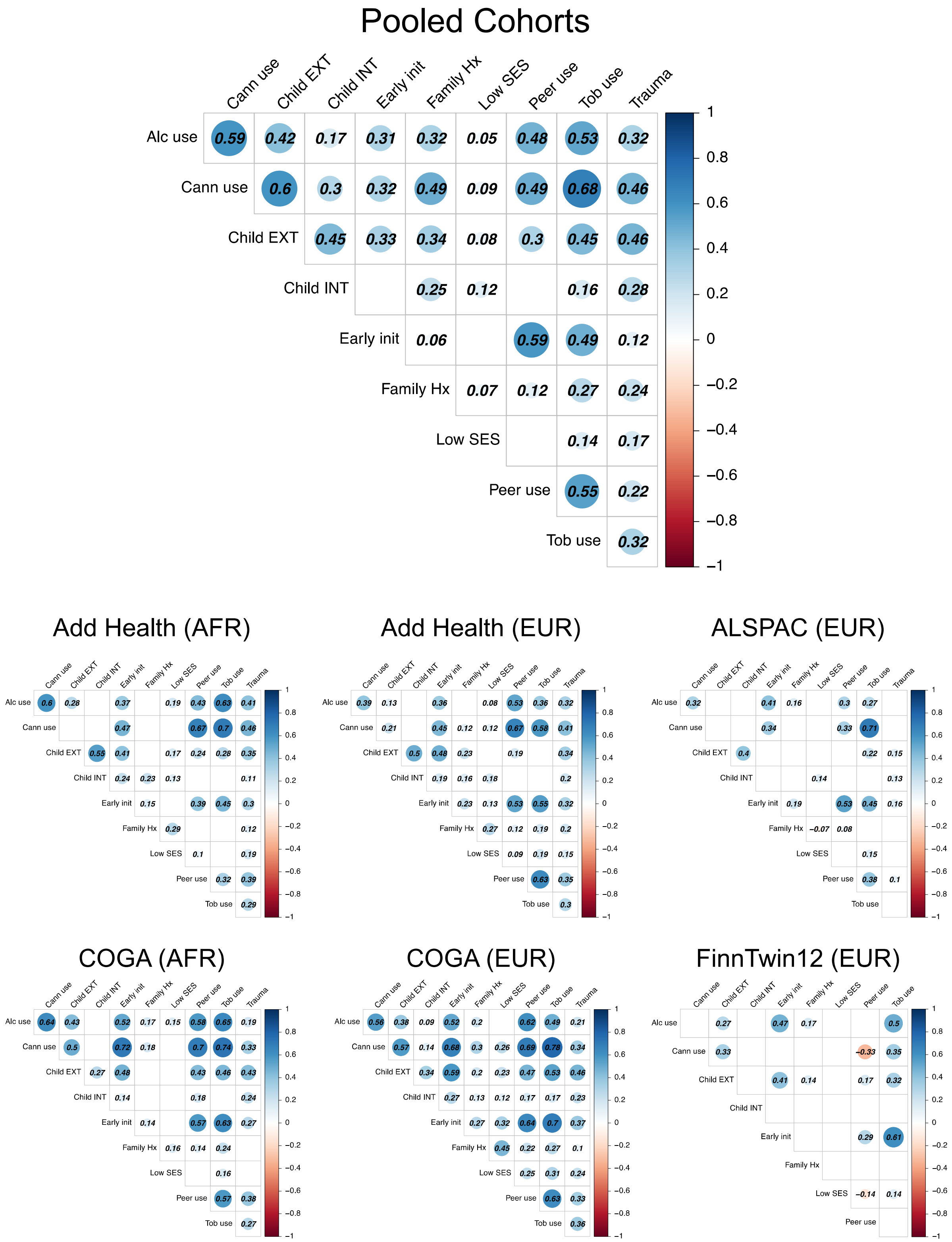

*Supplemental Figure 2: Tetrachoric Correlations Among Risk Index Items in Combined and Individual Cohorts*

### 4 GWAS selection and inclusion

We used summary statistics from recent genome wide association studies (GWAS) to create polygenic scores (PGS) in the four holdout samples. We chose GWAS for inclusion based on the fact that: 1) SUD show strong genetic overlap with other externalizing[27–29], internalizing[30,31], and psychotic disorders[32–34]; 2) both shared and substance specific genetic risk are associated with later SUDs[35–37]; 3) substance use and SUDs have only partial genetic overlap[38]; and 4) these samples had available results in both European and African ancestry cohorts.

#### 4.1 GWAS of externalizing (EXT)

Summary statistics used for EXT in the European ancestry cohorts come from the recent multivariate GWAS of externalizing problems by the Externalizing Consortium[39]. The Externalizing Consortium analyses focused on a GWAS of a latent factor for externalizing derived from seven input GWAS theorized to be part of the externalizing spectrum, including ADHD [40], problematic alcohol use [41,42], lifetime cannabis use [43], age of first sexual intercourse [44], number of sexual partners [44], general risk tolerance [44] and lifetime smoking initiation [45].These analyses converged onto a single factor. Polygenic scores for the latent externalizing factor were associated with externalizing factor scores in two holdout cohorts and with a variety of exploratory traits, including multiple substance use outcomes (both substance use and SUD).

For EXT in African ancestry cohorts, there is not an available multivariate GWAS that corresponds to the GWAS in European ancestries. Therefore, we performed a GWAS of an observed factor score in the COGA African ancestry cohort, derived from the same seven phenotypes used in the original Externalizing Consortium paper (and used for replication in the within family results in the European ancestry cohort). In order to ensure that there was no overlap between the discovery sample and COGA sample used in PGS analyses, we performed a ten-fold cross validation with leaving 10% of the sample out in every fold. GWAS from this analysis were used for PGS creation in the 10% not included in that run.

#### 4.2 GWAS of major depressive disorder (MDD)

Results for both the European and African ancestry GWAS come from a recent meta-analysis of large-scale major depressive disorder GWAS using data from the Psychiatric Genomics Consortium (PGC), UK Biobank (UKB), Million Veterans Program (MVP), FinnGen, and 23andMe [31]. While the original meta-analysis includes all of these samples (N ~1.2 million), we restricted the current analysis to the PGC, UKB, and MVP cohorts only in European ancestries (N ~720K) as we did not have access to the 23andMe data, and we wanted to eliminate the possibility of sample overlap between FinnGen and the FinnTwin12 sample. GWAS for the African ancestry cohorts come exclusively from the African ancestry results for MDD in MVP (N = 59,600).

#### 4.3 GWAS of problematic alcohol use (ALCP)

GWAS for problematic alcohol use (ALCP) in European ancestries is from a recent meta-analysis of GWAS for the PGC GWAS of alcohol dependence, the UKB GWAS of the problem subscale of the Alcohol Use Disorder Identification Test (AUDIT-P), and the MVP GWAS of alcohol use disorders (N ~ 430K)[32]. As Add Health, COGA, and FinnTwin12 were included in the original meta-analysis, we obtained GWAS results with each of those cohorts excluded for creating polygenic scores. Results for African ancestry come from the GWAS of AUD in MVP[46] (N ~ 56K).

#### 4.4 GWAS of alcohol consumption (ALCC)

We used results from the GWAS and Sequencing Consortium for Alcohol and Nicotine's (GSCAN) meta-analysis of drinks per week for alcohol consumption (ALCC) in European ancestries[45]. These results included the publicly available GSCAN results as well was the 23andMe data (N ~900K). Both ALSPAC and FinnTwin12 were included in the original meta-analysis, and we obtained GWAS results with each of those cohorts excluded. Results for African ancestry come from the GWAS of the consumption subscale of the Alcohol Use Disorder Identification Test (AUDIT-C) in MVP[46] (N ~ 56K).

#### 4.5 GWAS of schizophrenia (SCZ)

PGS for schizophrenia in the European ancestry cohorts were derived from the most recent iteration of the PGC's GWAS of SCZ (N ~130K)[47]. African ancestry results come from a meta-analysis of GWAS in the Genomic Psychiatry Cohort (GPC)[48] and Cooperative Studies Program (CSP) #572[49].

#### 4.6 GWAS of cigarettes per day/nicotine dependence (CPD)

For our smoking PGS in European ancestries, we used the publicly available GSCAN meta-analysis of cigarettes per day (CPD, N ~250K) [45]. These results again included ALSPAC and FinnTwin12, and we obtained GWAS results with each of those cohorts excluded. Results for PGS in African ancestries come from the most current GWAS of nicotine dependence [50] (N ~ 12K). While CPD and nicotine dependence are different phenotypes, the genetic correlation between the two is indistinguishable from one [50]. The GWAS of nicotine dependence included some COGA participants, and we obtained results with COGA excluded.

### 5 Polygenic Score Creation

##### 5.1.1 *Adjustment of GWAS effect sizes for linkage disequilibrium (LD)*

We adjusted GWAS effect sizes for the non-independence of nearby SNPs in the genome (referred to as linkage disequilibrium, or LD) using PRS-CSx[51], which employs a Bayesian continuous shrinkage parameter to correct for LD. We used ancestry matched samples from 1KG as a reference panel for both European (EUR) and African (AFR) ancestries.

Rather than using each of the target samples for the training sample, we utilized the 1KG ancestry matched samples and restricted to the ~1.3 million SNPs in the high-quality consensus genotype set defined by the HapMap 3 Consortium[52,53]. We generated polygenic scores using HapMap 3 SNPs that overlapped with the corresponding 1KG sample and UKB reference panel.

##### 5.1.2 *Polygenic scores*

We computed polygenic scores from the weighted sum of the effect-coded alleles for a given individual *i*:

$$S_i = \sum_{j=1}^{M} \hat{\beta}_j g_{ij}$$

where $S_i$ is the polygenic score, $\hat{\beta}_j$ is the estimated additive effect of the effect-coded allele at SNP $j$, and $g_{ij}$ is the genotype at SNP *j*. The polygenic scores were standardized within each study cohort. Because PRS-CSx improves predictive power for non-European ancestry samples with smaller GWAS, we utilized the “meta” option for the AFR ancestries, creating scores that were derived from the meta-analyzed EUR and AFR specific weights. In the European ancestries, we derived scores from the EUR weights alone (not meta-analyzed). In each cohort, this provided us with one PGS per phenotype in each cohort to carry forward include in the models for the pooled analyses.

To account for population stratification, we regressed each PGS on age, $age^2$, sex, sex*age, sex* $age^2$, and the first 10 ancestral PC’s. We then calculated the standardized residuals from these regression models for each of the six PGS (per cohort) and carried those forwards into the joint models that pooled the data from each cohort. Supplemental Figure 1, below, shows the GWAS matched for each PGS within each of the cohorts

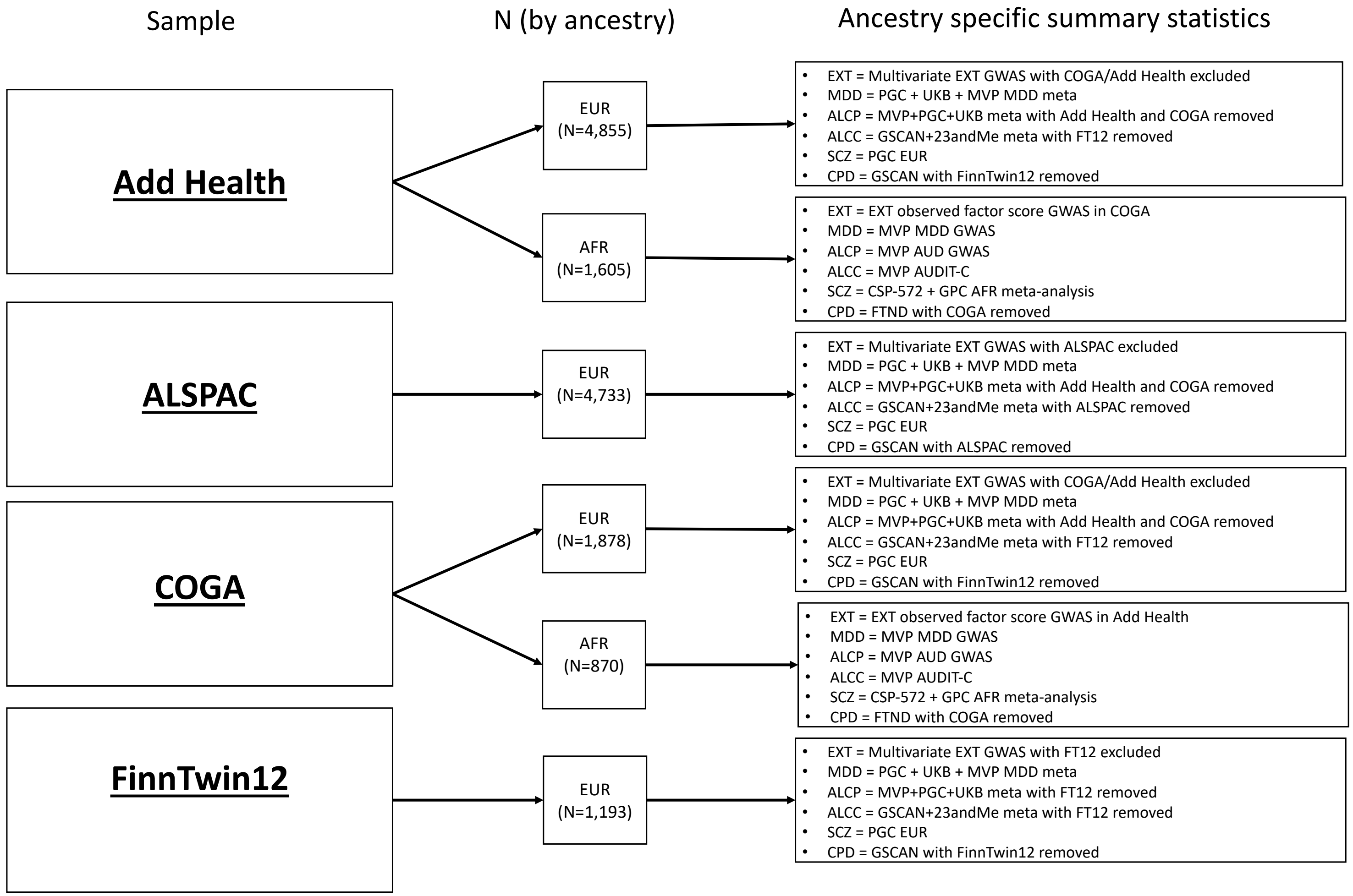


*Supplemental Figure 3: GWAS used for PGS creation in each cohort*

### 6 Deviations from preregistration

We made several important deviations from the preregistration that are worth noting, in the interest of transparency. These changes were added to the analysis plan, posted on the open science framework, along with date and time stamps.

For each of the changes from the original plan, our motivations were driven by ways to either improve the analysis or address a problem we did not foresee in the original preregistration.

(a) **Amendment (04/22/21):** The polygenic scores for major depressive disorder were expanded into a broader risk for internalizing after meta-analyzing with a GWAS of generalized anxiety disorder[57]. These GWAS showed relatively strong genetic overlap using bivariate LDSC[58] (rG ~.66).

(b) **Amendment (04/22/21):** We changed the PGS to those derived from PRS-CSx[51] (an extension of the original PRS-CS) as these allowed us to incorporate summary statistics from African ancestry GWAS and therefore create scores for the AFR subsamples in COGA and Add Health.

(c) **Amendment (04/22/21):** Due to issues with model convergence, we will use logistic regression in models with standard errors corrected for clustering at the family level[59].

(d) **Amendment (09/20/21):** Based on expert advice we will use an integrative data analysis approach[60] where we pool data and include cohort as a fixed effect. This approach is superior to meta-analysis because we have access to raw data.

(e) **Amendment (09/20/21):** We reverted to our original plan to use polygenic scores for major depressive disorder as a new GWAS with AFR ancestry results became available[31]. We will also include a polygenic score for schizophrenia based on the overlap between psychotic disorders and SUD[47], and the availability of ancestry matched results[48].

(f) **Amendment (09/28/21):** We changed our focus on SUDs from including both abuse and dependence to dependence only. This change was driven by the fact that some of the samples (specifically Add Health) had a large number of people meeting criteria for alcohol abuse, and the sample prevalence for AUD was particularly high (over 40%). We therefore used the more restrictive measure of dependence for each of the substances to ensure we were not incorrectly categorizing people as having an SUD when they do not. We also omitted count of substances for which people meet criteria for the sake of space (these models were never run).

(g) **Amendment (05/12/22):** We made the following changes based on requests from reviewers:

(i) Added nicotine dependence as its own independent outcome to fully cover the range of SUD phenotypes.

(ii) Included PGS for nicotine dependence/cigarettes per day [45,50] in addition to original PGS.

### 7 Variation in effect of clinical/environmental risk index (CERI)

To assess the relative impact of individual items, we ran a series of sensitivity analyses. The goal of these analyses was to ensure that the association between the CERI and each of the SUD phenotypes was not driven by any single item included in the CERI. We first estimated the association between individual risk factors and each of the SUD outcomes (Supplemental Figure 4). With the exception of the association between low childhood SES and alcohol dependence, each individual item is associated with increased odds of each of the SUD outcomes to varying degree. The one outlier for effect sizes of individual items was in regard to frequent adolescent cannabis use and both the drug dependence and any substance dependence outcomes.

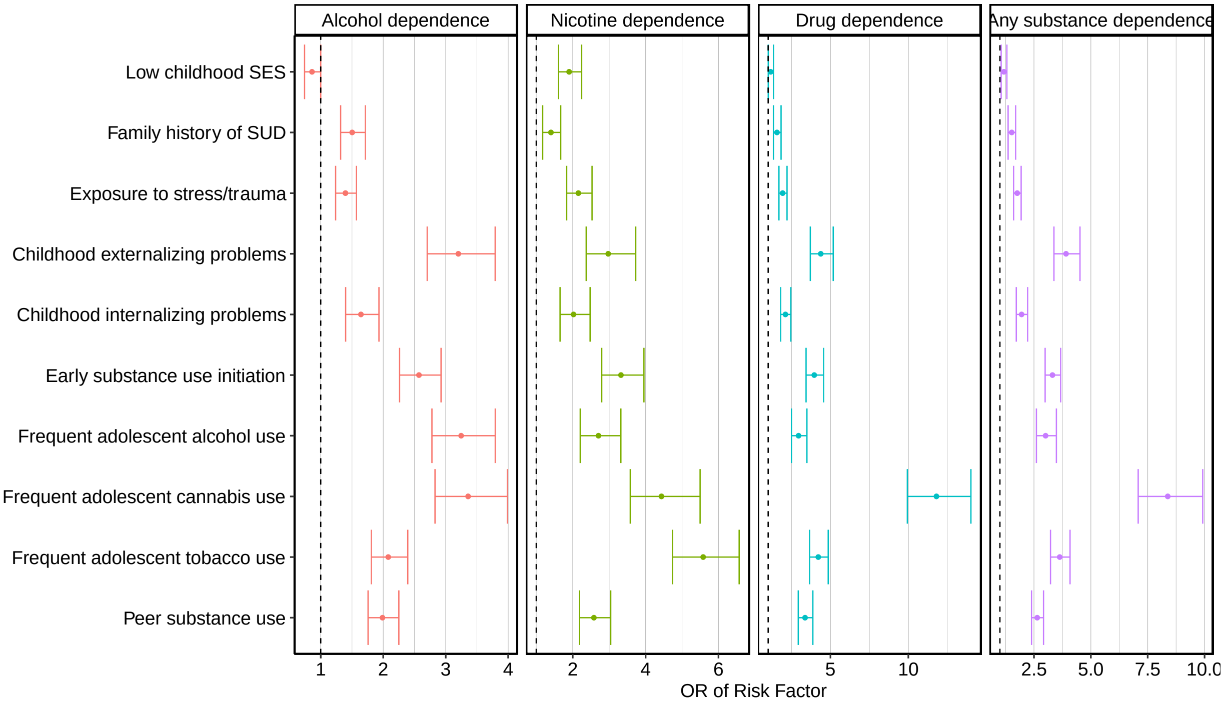


*Supplemental Figure 4: Associations between individual risk factors and SUDs*

In addition to testing the relative impact of each individual risk factor, we also evaluated the impact of removing one of the risk factors from the overall index to see relative change in the effect size. Supplemental Figure 5 presents the distribution of effect sizes for the CERI removing one of the risk factors for each of the SUD phenotypes. In each model, we also included sex, age, cohort, and all of the six PGSs (the Combined Risk Model). Panel A (Supplemental Figure 5) presents the distribution of the CERI effect sizes for each outcome. Overall, the effect sizes are relatively stable even when leaving one of the risk factors out. Panel B presents the same model, but with the removed risk factor included as a separate covariate. Again, the effect sizes are relatively stable, with two notable exceptions. The outlier for nicotine dependence is the effect size for the CERI when frequent adolescent tobacco use is included as a covariate. Similarly, the outlier for drug dependence is the effect size for the CERI when frequent adolescent cannabis use is included as a covariate. Even with these two outliers, to CERI is still significant and strongly associated with each SUD outcome.

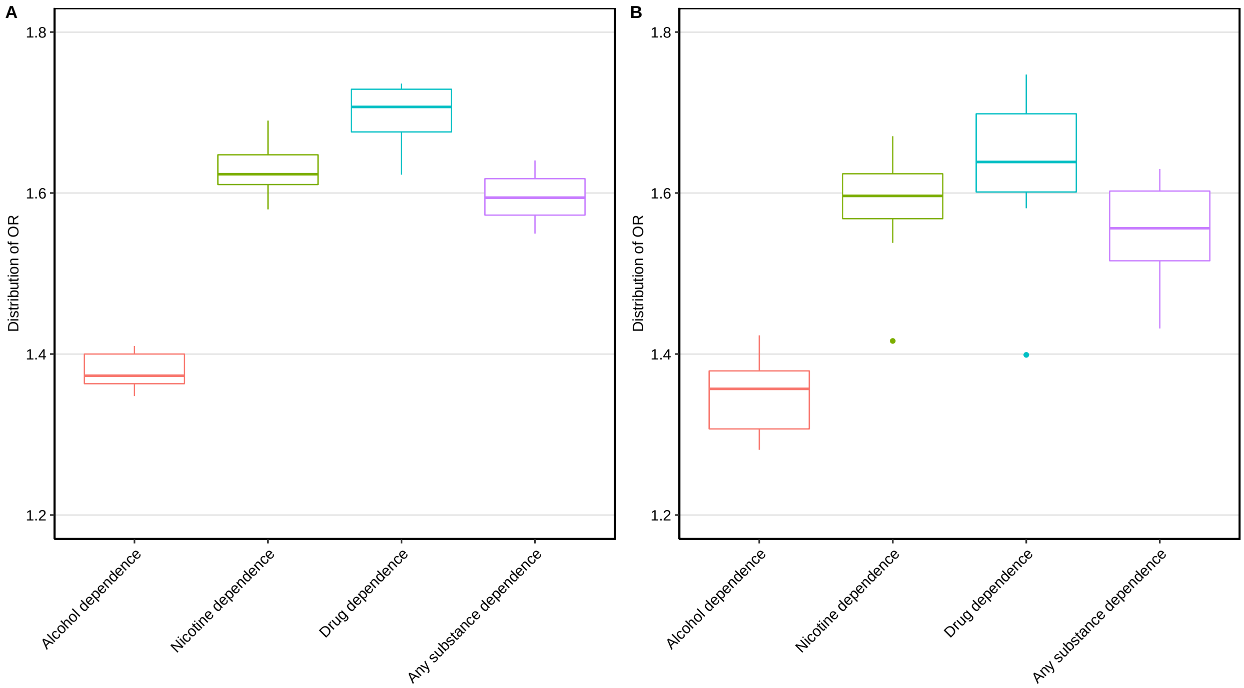


*Supplemental Figure 5: Effect Sizes for CERI with Individual Risk Factors Omitted*

#### 8 ROC Curves for CERI only and PGS only Models

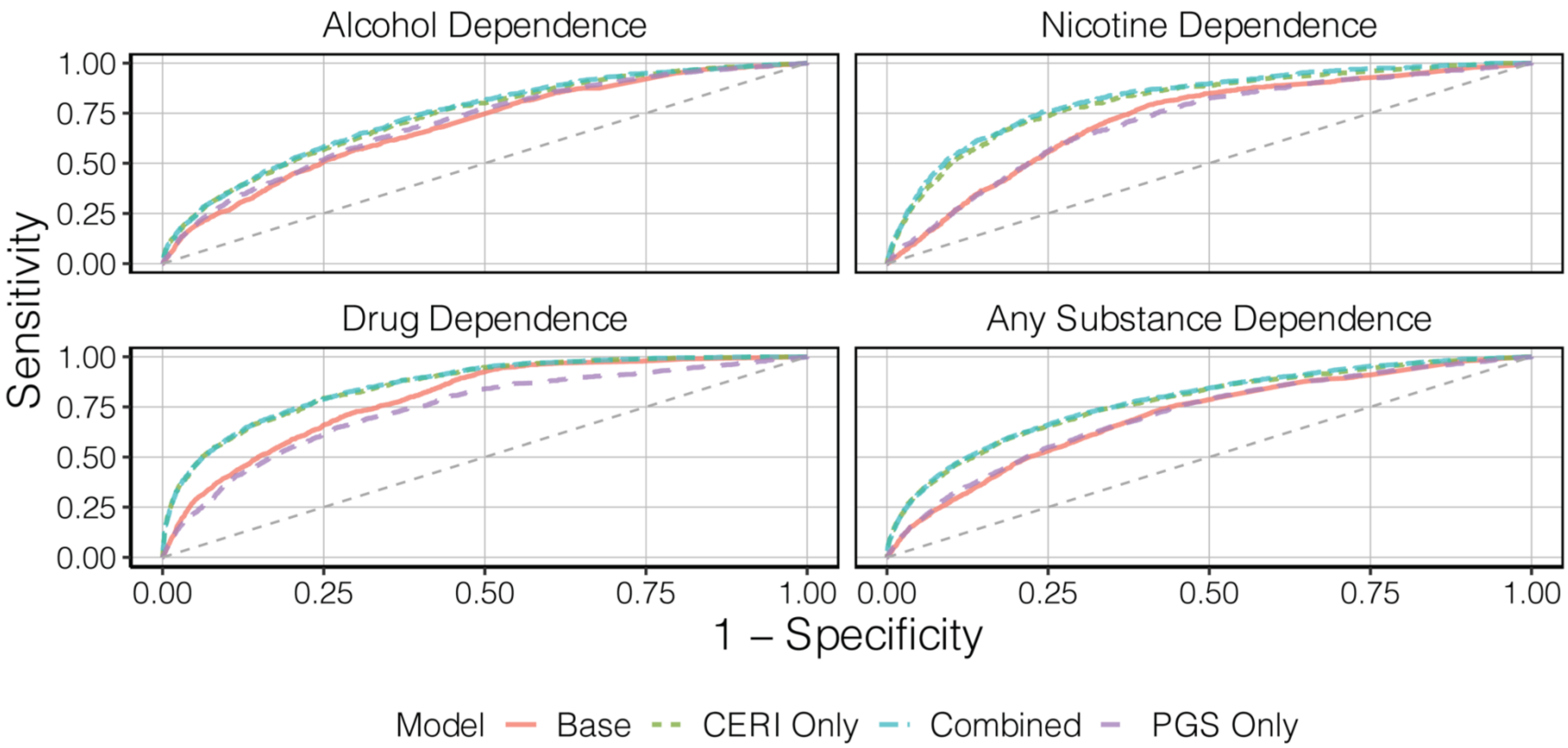


*Supplemental Figure 6: ROC Curves for Baseline (covariates only), CERI Only, PGS Only, and Combined Models*

AUC Estimates for Baseline, PGS, CERI, and Combined Models

| Phenotype | Model | AUC |
|---|---|---|
| Alcohol Dependence | Baseline (covariates only) | 0.688 |
| | CERI + covariates | 0.732 |
| | PGS + covariates | 0.701 |
| | Combined (PGS + CERI + covariates) | 0.738 |
| Nicotine Dependence | Baseline (covariates only) | 0.721 |
| | CERI + covariates | 0.811 |
| | PGS + covariates | 0.763 |
| | Combined (PGS + CERI + covariates) | 0.824 |
| Drug Dependence | Baseline (covariates only) | 0.793 |
| | CERI + covariates | 0.857 |
| | PGS + covariates | 0.806 |
| | Combined (PGS + CERI + covariates) | 0.860 |
| Any Substance Dependence | Baseline (covariates only) | 0.702 |
| | CERI + covariates | 0.772 |
| | PGS + covariates | 0.720 |
| | Combined (PGS + CERI + covariates) | 0.777 |

### 9 Random-effects Integrative Data Analysis (RE IDA)

In order to ensure the robustness of our results to and between-sample heterogeneity, we ran a complementary set of analyses alongside our fixed-effects (FE) IDA approach. The random effects (RE) approach assumes that the samples in the analysis represent random draws from a larger population distribution $\sim N(0, \sigma^2)$, as opposed to treating the effect of each cohort as known (and fixed, as in the fixed-effects approach). While there are more assumptions to the RE approach, the added advantages are that one can explicitly model between-study variation.

In our supplemental analyses, we tested for both random intercepts (for both study and family-unit) as well as testing for random slopes for each of the main predictors included in our analyses: the six polygenic scores (PGS) and the clinical/environmental risk index (CERI). In deciding the random-effects structure, we tested a series of nested models, adding random slopes and comparing the change in model fit using a $\chi^2$ difference test ($\chi^2_{Full}$ - $\chi^2_{Reduced}$). Once we identified the best fitting structure of the random effects, we estimated the models from the main analysis and compared the point estimates from the fixed effects and random effects models.

Supplemental Table 5 presents the tests for random slopes for the corresponding risk factors (6 PGS + CERI) with each of the SUD outcomes, compared to a baseline model which already includes a random intercept for cohort and family unit. We tested each random slope with each outcome, individually, as fitting all the random slopes at once was not possible. The model that included the random slope for the ALCC PGS showed improvement in overall fit above the baseline model in both alcohol dependence and any substance dependence. Likewise, for drug dependence, the model with a random slope for the EXT PGS showed significant improvement in fit. However, for each of the SUD outcomes, the biggest improvement in fit was gained by including a random slope for the CERI. We therefore included a random slope for the CERI, a random intercept for cohort, a random intercept for family unit, and a correlation between the random slope and the random intercept for cohort moving forward.

Supplemental Table 6 presents the parameter effects estimates from the models. Overwhelmingly, the parameter estimates from the random effects IDA approach, which explicitly models the between sample heterogeneity in the effect of the CERI, were consistent with the results from the main analysis (e.g., the fixed-effects IDA). Overall, these results support the findings from the main analyses and demonstrate that between-sample heterogeneity is not the reason for the associations between either the CERI or PGSs and each of SUD outcomes.

#### 10 References


1. Degenhardt, L. *et al.* The global burden of disease attributable to alcohol and drug use in 195 countries and territories, 1990–2016: a systematic analysis for the Global Burden of Disease Study 2016. *The Lancet Psychiatry* **5**, 987–1012 (2018).
2. Sacks, J. J., Gonzales, K. R., Bouchery, E. E., Tomedi, L. E. & Brewer, R. D. 2010 National and State Costs of Excessive Alcohol Consumption. *American Journal of Preventive Medicine* **49**, e73–e79 (2015).
3. National Drug Intelligence Center. *National drug threat assessment*. vol. 2019 (2011).
4. Florence, C. S., Zhou, C., Luo, F. & Xu, L. The Economic Burden of Prescription Opioid Overdose, Abuse, and Dependence in the United States, 2013. *Med Care* **54**, 901–906 (2016).
5. Verhulst, B., Neale, M. C. & Kendler, K. S. The heritability of alcohol use disorders: a meta-analysis of twin and adoption studies. *Psychol Med* **45**, 1061–1072 (2015).
6. Verweij, K. J. H. *et al.* Genetic and environmental influences on cannabis use initiation and problematic use: A meta-analysis of twin studies. *Addiction* **105**, 417–430 (2010).
7. Burt, S. A. Are there meaningful etiological differences within antisocial behavior? Results of a meta-analysis. *Clin Psychol Rev* **29**, 163–178 (2009).
8. Rhee, S. H. & Waldman, I. D. Genetic and environmental influences on antisocial behavior: A meta-analysis of twin and adoption studies. *Psychol Bull* **128**, 490–529 (2002).
9. Kendler, K. S., Jacobson, K. C., Prescott, C. A. & Neale, M. C. Specificity of Genetic and Environmental Risk Factors for Use and Abuse/Dependence of Cannabis, Cocaine, Hallucinogens, Sedatives, Stimulants, and Opiates in Male Twins. *American Journal of Psychiatry* **160**, 687–695 (2003).
10. Meier, M. H. *et al.* Which adolescents develop persistent substance dependence in adulthood? Using population-representative longitudinal data to inform universal risk assessment. *Psychol Med* **46**, 877–889 (2016).
11. Barr, P. B. *et al.* Using polygenic scores for identifying individuals at increased risk of substance use disorders in clinical and population samples. *Translational Psychiatry* **10**, 196 (2020).
12. Gu, F. *et al.* Combining common genetic variants and non-genetic risk factors to predict risk of cutaneous melanoma. *Human Molecular Genetics* (2018) doi:10.1093/hmg/ddy282.
13. O'Sullivan, J. W. *et al.* Combining clinical and polygenic risk improves stroke prediction among individuals with atrial fibrillation. *medRxiv* 2020.06.17.20134163 (2020) doi:10.1101/2020.06.17.20134163.
14. Harris, K. M., Halpern, C. T., Haberstick, B. C. & Smolen, A. The National Longitudinal Study of Adolescent Health (Add Health) sibling pairs data. *Twin Research and Human Genetics* **16**, 391–398 (2013).
15. The Haplotype Reference Consortium *et al.* A reference panel of 64,976 haplotypes for genotype imputation. *Nature Genetics* **48**, 1279–1283 (2016).
16. Auton, A. *et al.* A global reference for human genetic variation. *Nature* **526**, 68–74 (2015).
17. Boyd, A. *et al.* Cohort profile: The 'Children of the 90s'-The index offspring of the avon longitudinal study of parents and children. *International Journal of Epidemiology* (2013) doi:10.1093/ije/dys064.
18. Fraser, A. *et al.* Cohort profile: The avon longitudinal study of parents and children: ALSPAC mothers cohort. *International Journal of Epidemiology* **42**, 97–110 (2013).
19. Northstone, K. *et al.* The Avon Longitudinal Study of Parents and Children (ALSPAC): an update on the enrolled sample of index children in 2019 [version 1; peer review: 2 approved]. *Wellcome Open Research* **4**, (2019).

20. Harris, P. A. *et al.* Research electronic data capture (REDCap)-A metadata-driven methodology and workflow process for providing translational research informatics support. *Journal of Biomedical Informatics* **42**, 377–381 (2009).
21. Taylor, A. E. *et al.* Exploring the association of genetic factors with participation in the Avon Longitudinal Study of Parents and Children. *International Journal of Epidemiology* **47**, dyy060 (2018).
22. Edenberg, H. J. The collaborative study on the genetics of alcoholism: An update. *Alcohol Research and Health* **26**, 214–218 (2002).
23. Bucholz, K. K. *et al.* A new, semi-structured psychiatric interview for use in genetic linkage studies: a report on the reliability of the SSAGA. *Journal of Studies on Alcohol* **55**, 149–158 (1994).
24. Bucholz, K. K. *et al.* Comparison of Parent, Peer, Psychiatric, and Cannabis Use Influences Across Stages of Offspring Alcohol Involvement: Evidence from the COGA Prospective Study. *Alcoholism: Clinical and Experimental Research* **41**, 359–368 (2017).
25. Rose, R. J. R. J. *et al. FinnTwin12 Cohort: An Updated Review*. *Twin Research and Human Genetics* vol. 22 (2019).
26. Kaprio, J. The Finnish Twin Cohort Study: an update. *Twin Res Hum Genet* **16**, 157–162 (2013).
27. Barr, P. B. & Dick, D. M. The Genetics of Externalizing Problems. *Current Topics in Behavioral Neurosciences* **47**, 93–112 (2020).
28. Krueger, R. F. *et al.* Etiological connections among substance dependence, antisocial behavior and personality: Modeling the externalizing spectrum. *J Abnorm Psychol* **111**, 411–424 (2002).
29. Kendler, K. S. & Myers, J. The boundaries of the internalizing and externalizing genetic spectra in men and women. *Psychological Medicine* **44**, 647–655 (2014).
30. Polimanti, R. *et al.* Evidence of causal effect of major depression on alcohol dependence: Findings from the psychiatric genomics consortium. *Psychological Medicine* (2019) doi:10.1017/S0033291719000667.
31. Levey, D. F. *et al.* Bi-ancestral depression GWAS in the Million Veteran Program and meta-analysis in >1.2 million individuals highlight new therapeutic directions. *Nature Neuroscience* (2021) doi:10.1038/s41593-021-00860-2.
32. Zhou, H. *et al.* Genome-wide meta-analysis of problematic alcohol use in 435,563 individuals yields insights into biology and relationships with other traits. *Nature Neuroscience* (2020) doi:10.1038/s41593-020-0643-5.
33. Johnson, E. C. *et al.* A large-scale genome-wide association study meta-analysis of cannabis use disorder. *The Lancet Psychiatry* (2020) doi:10.1016/S2215-0366(20)30339-4.
34. Zhou, H. *et al.* Association of OPRM1 Functional Coding Variant With Opioid Use Disorder: A Genome-Wide Association Study. *JAMA Psychiatry* (2020) doi:10.1001/jamapsychiatry.2020.1206.
35. Kendler, K. S., Gardner, C. & Dick, D. M. Predicting alcohol consumption in adolescence from alcohol- specific and general externalizing genetic risk factors, key environmental exposures and their interaction. *Psychol Med* **41**, 1507–1516 (2011).
36. Meyers, J. L. *et al.* Genetic Influences on Alcohol Use Behaviors Have Diverging Developmental Trajectories : A Prospective Study Among Male and Female Twins. *Alcoholism: Clinical and Experimental Research* **38**, 2869–2877 (2014).
37. Barr, P. B. *et al.* Parsing Genetically Influenced Risk Pathways: Genetic Loci Impact Problematic Alcohol Use Via Externalizing and Specific Risk. *medRxiv* 2021.07.20.21260861 (2021) doi:10.1101/2021.07.20.21260861.

38. Sanchez-Roige, S., Palmer, A. A. & Clarke, T. K. Recent Efforts to Dissect the Genetic Basis of Alcohol Use and Abuse. *Biological Psychiatry* (2020) doi:10.1016/j.biopsych.2019.09.011.
39. Karlsson Linnér, R. *et al.* Multivariate analysis of 1.5 million people identifies genetic associations with traits related to self-regulation and addiction. *Nature Neuroscience* 1–10 (2021) doi:10.1038/s41593-021-00908-3.
40. Demontis, D. *et al.* Discovery of the first genome-wide significant risk loci for attention deficit/hyperactivity disorder. *Nature Genetics* **51**, 63–75 (2019).
41. Walters, R. K. *et al.* Trans-ancestral GWAS of alcohol dependence reveals common genetic underpinnings with psychiatric disorders. *Nature Neuroscience* **21**, 1656–1669 (2018).
42. Sanchez-Roige, S. *et al.* Genome-wide association study meta-analysis of the alcohol use disorders identification test (AUDIT) in two population-based cohorts. *American Journal of Psychiatry* **176**, 107–118 (2019).
43. Pasman, J. A. *et al.* GWAS of lifetime cannabis use reveals new risk loci, genetic overlap with psychiatric traits, and a causal influence of schizophrenia. *Nature Neuroscience* **21**, 1161–1170 (2018).
44. Karlsson Linnér, R. *et al.* Genome-wide association analyses of risk tolerance and risky behaviors in over 1 million individuals identify hundreds of loci and shared genetic influences. *Nature Genetics* **51**, 245–257 (2019).
45. Liu, M. *et al.* Association studies of up to 1.2 million individuals yield new insights into the genetic etiology of tobacco and alcohol use. *Nature Genetics* **51**, 237–244 (2019).
46. Kranzler, H. R. *et al.* Genome-wide association study of alcohol consumption and use disorder in 274,424 individuals from multiple populations. *Nature Communications* **10**, 1499 (2019).
47. Trubetskoy, V. *et al.* Mapping genomic loci implicates genes and synaptic biology in schizophrenia. *Nature 2022* 1–13 (2022) doi:10.1038/s41586-022-04434-5.
48. Bigdeli, T. B. *et al.* Contributions of common genetic variants to risk of schizophrenia among individuals of African and Latino ancestry. *Mol Psychiatry* (2019) doi:10.1038/s41380-019-0517-y.
49. Bigdeli, T. B. *et al.* Genome-Wide Association Studies of Schizophrenia and Bipolar Disorder in a Diverse Cohort of US Veterans. *Schizophrenia Bulletin* (2020) doi:10.1093/schbul/sbaa133.
50. Quach, B. C. *et al.* Expanding the genetic architecture of nicotine dependence and its shared genetics with multiple traits. *Nature Communications* **11**, (2020).
51. Ruan, Y. *et al.* Improving Polygenic Prediction in Ancestrally Diverse Populations. *Nature Genetics* (2022) doi:10.1038/s41588-022-01054-7.
52. Altshuler, D. M. *et al.* Integrating common and rare genetic variation in diverse human populations. *Nature* **467**, 52–58 (2010).
53. Ge, T., Chen, C.-Y., Ni, Y., Feng, Y.-C. A. & Smoller, J. W. Polygenic prediction via Bayesian regression and continuous shrinkage priors. *Nature Communications* **10**, 1776 (2019).
54. Sher, K. J., Grekin, E. R. & Williams, N. A. The development of alcohol use disorders. *Annual Review of Clinical Psychology* **1**, 493–523 (2005).
55. Hughes, K. *et al.* The effect of multiple adverse childhood experiences on health: a systematic review and meta-analysis. *The Lancet Public Health* (2017) doi:10.1016/S2468-2667(17)30118-4.
56. McCutcheon, V. V. *et al.* Familial association of abstinent remission from alcohol use disorder in first-degree relatives of alcohol-dependent treatment-seeking probands. *Addiction* **112**, 1909–1917 (2017).

57. Levey, D. F. *et al.* Reproducible Genetic Risk Loci for Anxiety: Results From ~200,000 Participants in the Million Veteran Program. *Am J Psychiatry* (2020) doi:10.1176/appi.ajp.2019.19030256.
58. Bulik-Sullivan, B. K. *et al.* LD Score regression distinguishes confounding from polygenicity in genome-wide association studies. *Nature Genetics* **47**, 291–295 (2015).
59. Zeileis, A., Köll, S. & Graham, N. Various versatile variances: An object-oriented implementation of clustered covariances in r. *Journal of Statistical Software* **95**, 1–36 (2020).
60. Curran, P. J. & Hussong, A. M. Integrative Data Analysis: The Simultaneous Analysis of Multiple Data Sets. *Psychological Methods* **14**, 81–100 (2009).
